# Cardiac impairment in Long Covid 1-year post-SARS-CoV-2 infection

**DOI:** 10.1101/2022.04.03.22272610

**Authors:** Adriana Roca-Fernández, Malgorzata Wamil, Alison Telford, Valentina Carapella, Alessandra Borlotti, David Monteiro, Helena Thomaides-Brears, Matthew D Kelly, Andrea Dennis, Rajarshi Banerjee, Matthew D. Robson, Michael Brady, Gregory Y. H. Lip, Sacha Bull, Melissa Heightman, Ntobeko Ntusi, Amitava Banerjee

## Abstract

**Background:** Long Covid is associated with multiple symptoms and impairment in multiple organs. Cardiac impairment has been reported to varying degrees by varying methodologies in cross-sectional studies. Using cardiac magnetic resonance (CMR), we investigated the 12-month trajectory of cardiac impairment in individuals with Long Covid.

**Methods:** 534 individuals with Long Covid underwent baseline CMR (T1 and T2 mapping, cardiac mass, volumes, function, and strain) and multi-organ MRI at 6 months (IQR 4.3,7.3) since first post-COVID-19 symptoms and 330 were rescanned at 12.6 (IQR 11.4, 14.2) months if abnormal findings were reported at baseline. Symptoms, standardised questionnaires, and blood samples were collected at both timepoints. Cardiac impairment was defined as one or more of: low left or right ventricular ejection fraction (LVEF and RVEF), high left or right ventricular end diastolic volume (LVEDV and RVEDV), low 3D left ventricular global longitudinal strain (GLS), or elevated native T1 in ≥3 cardiac segments. A significant change over time was reported by comparison with 92 healthy controls.

**Results:** The technical success of this multiorgan assessment in non-acute settings was 99.1% at baseline, and 98.3% at follow up, with 99.6% and 98.8% for CMR respectively. Of individuals with Long Covid, 102/534 [19%] had cardiac impairment at baseline; 71/102 had complete paired data at 12 months. Of those, 58% presented with ongoing cardiac impairment at 12 months. High sensitivity cardiac troponin I and B-type natriuretic peptide were not predictive of CMR findings, symptoms, or clinical outcomes. At baseline, low LVEF, high RVEDV and low GLS were associated with cardiac impairment. Low LVEF at baseline was associated with persistent cardiac impairment at 12 months.

**Conclusion:** Cardiac impairment, other than myocarditis, is present in 1 in 5 individuals with Long Covid at 6 months, persisting in over half of those at 12 months. Cardiac-related blood biomarkers are unable to identify cardiac impairment in Long COVID. Subtypes of disease (based on symptoms, examination, and investigations) and predictive biomarkers are yet to be established. Interventional trials with pre-specified subgroup analyses are required to inform therapeutic options.

## Introduction

Cardiovascular disease (CVD), the commonest cause of global morbidity and mortality, has been linked with COVID-19 severity and mortality since the first reports from Wuhan in early 2020 (1–3). The pandemic has not only caused direct COVID-19-related effects in those with CVD; there have also been profound indirect effects, particularly due to disruption of CVD services (referral, diagnosis, and treatment) (4–6). Underlying CVD may also increase risk of longer-term symptoms, admission, and mortality in individuals recovering from COVID-19 hospitalization, which is associated with increased risk of multi-organ (including cardiac) impairment, compared to the general population (7). Moreover, there is evidence of multi-organ impairment in non-hospitalised COVID-19 (8–10).

Long COVID, classified as persistent symptoms ≥12 weeks following SARS-CoV-2 infection, is still being defined whether by symptoms, signs, organ impairment or recovery. Dyspnoea, palpitations and chest pain are common, even 1 year after infection (11), but associations with cardiac impairment are unclear, and the subtypes more likely to recover have not been identified. In a large UK, post-COVID assessment service, almost half of individuals where cardiac magnetic resonance (CMR) scans were performed, had evidence of mild myocarditis (12) and symptom improvement at 6 months was neither correlated with improvement on CMR imaging nor lung parenchymal recovery (13).

A systematic review of CMR findings post-COVID-19 identified myocarditis as the most prevalent diagnosis (40%) (14), T1 abnormalities and oedema on T2 as the most common findings, and occasional late gadolinium enhancement (LGE). These findings may be present even in absence of elevated cardiac blood biomarkers (e.g. troponin or NT-pro-BNP, B-type natriuretic peptide) (14,15). Pericardial effusion and reduced LV and RV function have been occasionally reported, but pericarditis is rare. Due to heterogeneity of study designs, patient populations, clinical pathways and access to services, cardiac impairment in Long COVID at baseline and over time are ill-defined (15).

Although echocardiography is often the first choice for assessment of cardiac function, CMR is the gold-standard assessment, with lower inter- and intra-observer variability, ensuring a more accurate assessment of cardiac structure and function. We therefore conducted a prospective, longitudinal 1-year study in individuals with Long COVID to investigate: 1) characteristics and trajectory of cardiac impairment; 2) impact of acute hospitalisation for COVID-19 on cardiac impairment; and 3) design of clinical management pathways for individuals at risk of cardiac impairment.

## Methods

### Population and study design

The COVERSCAN study (NCT04369807) is a prospective, ongoing study of organ function using quantitative MRI in individuals with persistent COVID-19 symptoms. Individuals were recruited via advertisement in Long COVID support groups and invited to undergo CoverScan (Perspectum, Oxford, UK), a multiparametric MRI assessment of lungs, heart, liver, pancreas, kidneys, and spleen. Assessment was at Perspectum (Oxford), Mayo Clinic (London) and Chenies Mews Imaging Centre (London), between May 2020 and August 2021 **(Figure 1)**. Healthy controls were recruited within the same period and scanned twice on the same date to assess repeatability. COVID-19 was classified by either laboratory-confirmed SARS-CoV-2 infection (159 tested SARS-CoV-2-positive by oropharyngeal/nasopharyngeal swab for reverse-transcriptase PCR; 150 individuals with positive antibodies) or strong clinical suspicion of SARS-CoV-2 infection with typical symptoms/signs (245 individuals). Exclusion criteria were symptoms of active respiratory viral infection (temperature >37.8°C or ≥3 episodes of coughing in 24 hours), hospital discharge in the last 7 days, and contraindications to MRI, including implanted pacemakers, defibrillators, other metallic implanted devices, and claustrophobia. Participants gave written informed consent. Those with organ impairment at baseline MRI scan (in ≥1 of the following organs: lungs, heart, liver, pancreas, spleen, kidneys) or blood tests were invited back for 6-month follow-up. Incidental findings classified as benign and/or not requiring follow-up by an experienced radiologist were not invited for follow-up.

**Figure 1:**
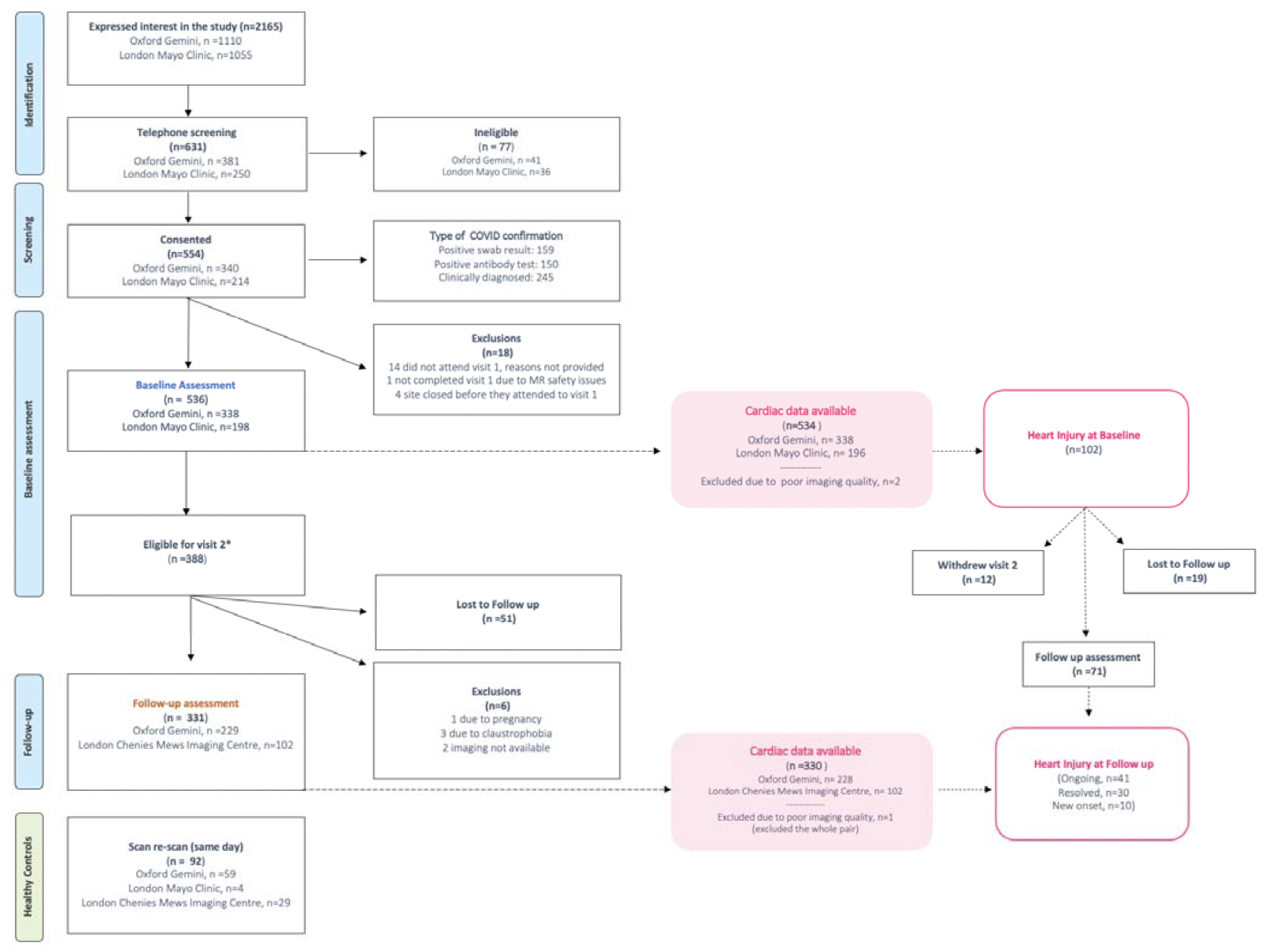
Study population for cardiac complications of Long COVID. * Individuals were eligible for follow up when MRI impairment or abnormal bloods in any organ were found at baseline.

### Symptoms, quality of life and function

Presence and severity of symptoms were assessed by self-report and validated questionnaires: EQ-5D-5L (Utility score and quality of life related to usual activities), and Dyspnoea-12 at baseline and follow-up, when Left Ventricular Function Questionnaire (LVD-36) was also conducted (**Supplementary methods 1**). Time off work due to Long COVID was recorded as total number of days at follow up.

### Blood investigations

Two blood samples were taken at both timepoints, on the same day as the MRI scan: one immediately sent for analysis, the other fractionated and frozen for later analysis **(Supplementary methods 2)**.

### Multiorgan imaging

Participants were scanned at Perspectum Gemini (Oxford: n=338; MAGNETOM Aera 1.5T scanner) and Mayo Clinic (London: n=198; MAGNETOM Vida 3T) (both scanners: Siemens Healthcare, Erlangen, Germany), at baseline and follow-up with multi-organ, multi-parametric MRI assessment (total ∼40min duration). All imaging methods were deployed in standard clinical MRI scanners as expedited versions of previously published methods, which unless otherwise stated, utilised short (<14seconds) breath-holds **(Supplementary methods 3, 4)**. To evaluate measurement repeatability, two separate scans were performed in healthy controls (1.5T, n=59; 3T, n=33) on the same day. After first scan, the participant had a 10-minute break out of the scanner before a second identical scan. Technical success was by reporting quality-assured measures for each variable, and overall, in report delivery for each patient.

#### Reference ranges

Reference ranges were from 92 healthy, sex- and age-matched controls **(Supplementary methods 5, Table S1)**. For each metric, we computed 2.5% and 97.5% percentiles using bootstrapping (100,000 permutations), except pancreas proton-density fat fraction (PDFF), where the 95% percentile was for the upper limit, and liver cT1 and PDFF, where we used established thresholds(27). Reference ranges for organ length and volume required larger sample size for sex- and height-stratification, so we used a sample of 1836 individuals from UK Biobank without self-reported diabetes or hypertension.

#### Definition of cardiac and multi-organ impairment

Beyond myocardial changes, cardiac impairment was defined by consensus as ≥1 of the following outside reference range: left or right ventricular ejection fraction (LVEF or RVEF) or left or right ventricular end diastolic volume (LVEDV or RVEDV), 3D GLS (abnormal will be referred as low, in absolute values) or ≥3 quantitative T1 mapping segments. Multi-organ impairment was defined as ≥2 measurements outside reference ranges in a further organ (excluding elevated liver or kidney volume)(17)(Further details in **supplementary methods 5, Table S1)**.

#### Community-delivered diagnostic assessment

Technical success of CMR was determined by reporting quality-assured measures for each variable reported herein, and overall, in delivering a report for each patient. For cardiac T1 and T2, technical success was based on value availability for least 3 AHA segments. Clinical utility was not assessed.

#### Statistical Analysis

We used R software version 4.0.4 and p-values <0.05 defined statistically significance. For parametric and non-parametric continuous, and categorical variables, we used mean (standard deviation, SD) and median (interquartile range, IQR), and frequency (percentage), respectively. For group-wise comparisons of continuous parametric and non-parametric, and categorical variables, T-test, Wilcoxon rank sum and Fisher’s exact tests, respectively, were used. Repeatability coefficients (RC) for each CMR metric in healthy controls determined the smallest detectable difference between repeated measures(18). Change from baseline was assumed only when change over time was greater than the RC. Baseline and follow-up metrics were assessed using reference ranges calculated in healthy controls. Associations with all exposures were by logistic and linear regression for categoric and continuous dependent variables, respectively. Multivariate stepwise regressions were performed to assess which explanatory variables were most predictive.

## Results

### Study population and symptoms

Of 536 individuals enrolled at baseline, 534 had available CMR data at a median 6 (IQR (4.33,7.26)) months after first COVID-19 symptoms (**Table 1, Figure 1**). Of those, 6 (1%) presented with raised cardiac blood biomarkers (high hs-cTnI, n=4 and high NT-proBNP, n= 2), but only 1/6 had abnormal CMR with both low LVEF and RVEF at 6 months and acute COVID-19 hospitalization. Another 101 individuals (19%) had cardiac impairment on CMR with normal cardiac blood biomarkers **(Figure 2)**. At 6 months, 62/102 (62%) individuals with cardiac impairment had severe Long COVID, based on questionnaires **(S1)**. Forty-three (43%) and 44 (44%) individuals had severe and moderate symptoms respectively; most commonly fatigue (100%), shortness of breath (88%), headache (83%), chest pain (81%) and cough (80%). Symptom prevalence was similar regardless of cardiac impairment **(Table 1)**.

**Table 1:**
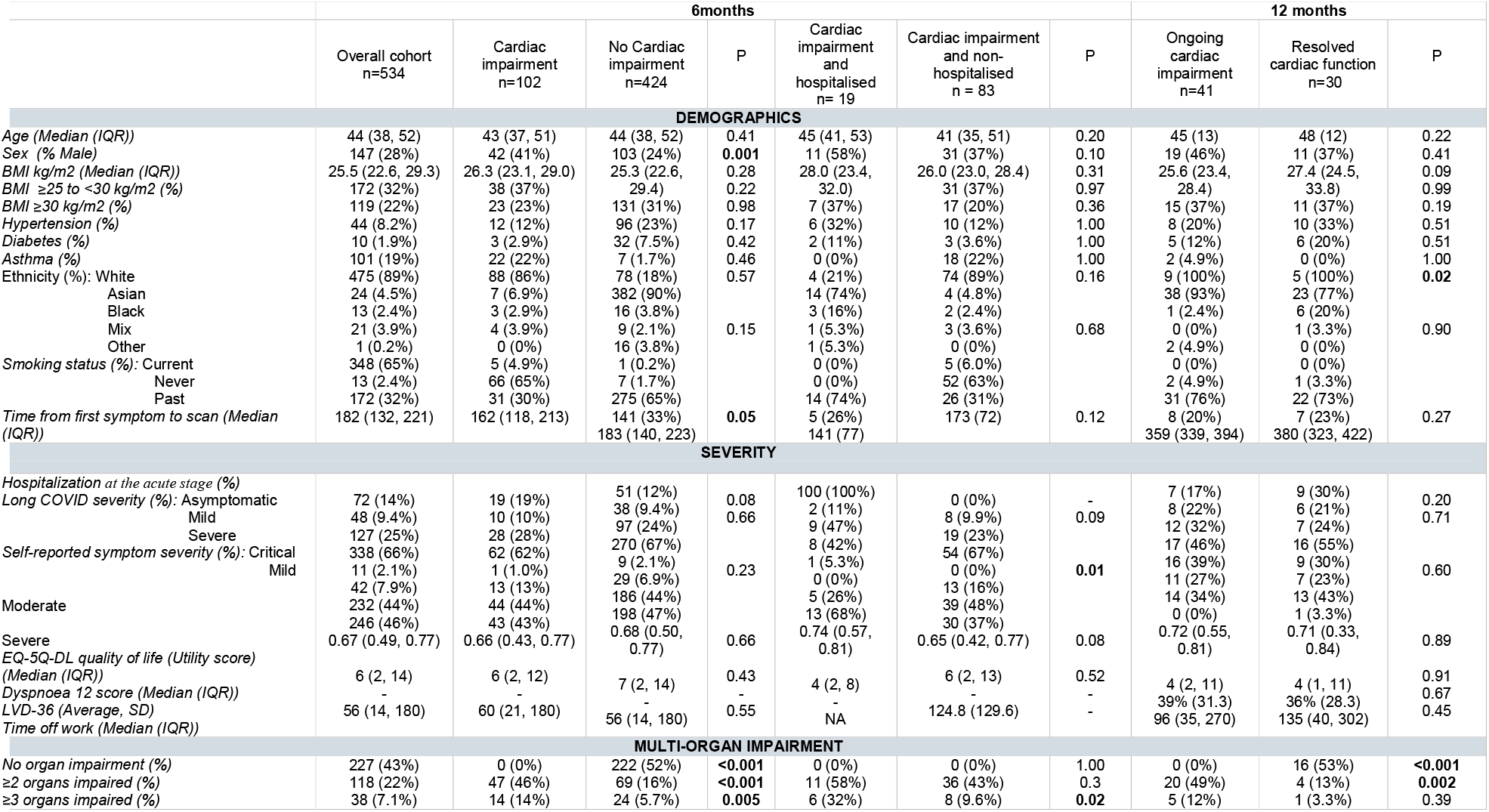
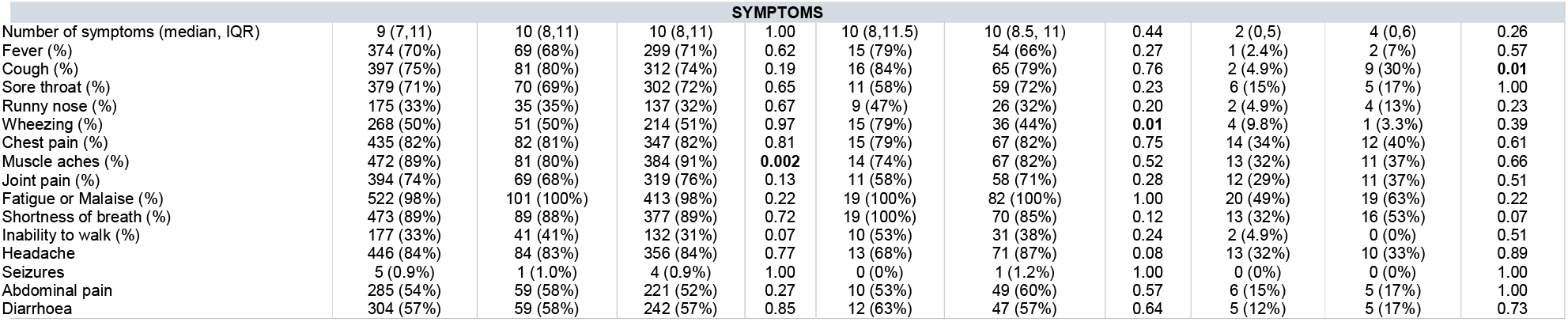
Characteristics for overall population, cardiac impairment vs no cardiac impairment at 6 and 12 months in individuals with Long Covid.

**Figure 2:**
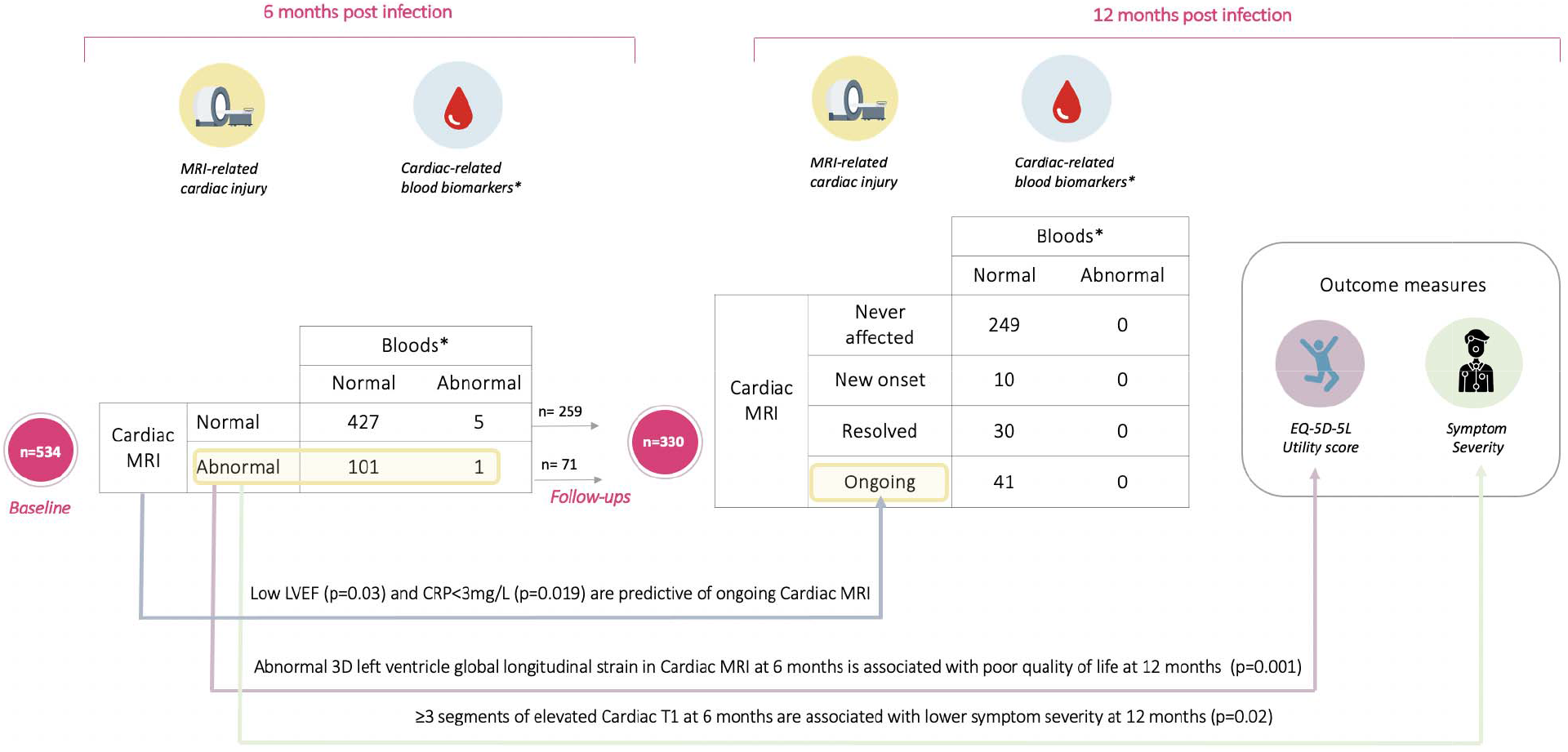
Evolution and characteristics of cardiac impairment in Long Covid 1-year post-SARS-CoV-2 infection. Numbers in the table are referring to number of patients. Abbreviations: LVEF, left ventricular ejection fraction; MRI, magnetic resonance imaging; CRP, C-reactive protein; mg/L, milligrams per litre. *Referring to High sensitivity cardiac troponin I (hs-cTnI) and B-type natriuretic peptide (NT-proBNP)

### CMR, organ impairment and blood investigations

Technical success of CMR and integrated assessment was 99.6% at first visit. Cardiac impairment was mostly characterized by reduced LVEF (21/102, 21%) or RVEF (21/102, 21%), low GLS (21/102, 21%) or T1 findings (46/102, 45%) **(Table 2)**. Multi-organ involvement (≥3 organs) was more common in those with versus without cardiac impairment (14% vs 5.7%, p=0.005) (**Table 1**). Only 1 individual with cardiac impairment on CMR had abnormal NT-proBNP. Abnormal MCHC (p=0.04), chloride (p=0.05) and amylase (p=0.03) levels were more frequent in cardiac impairment **(Table S2)**. No blood investigations were predictive of organ impairment at 6 months.

**Table 2:** Cardiac impairment by abnormal cardiac magnetic resonance findings at 6 and 12 months in individuals with Long Covid.

|  | 6 months |  |  |  |  |  | 12 months |  |  |
| --- | --- | --- | --- | --- | --- | --- | --- | --- | --- |
|  | Cardiac impairment<br>n=102 | No Cardiac impairment<br>n=424 | P | Cardiac impairment and hospitalised,<br>n = 19 | Cardiac impairment not hospitalised,<br>n = 83 | P | Ongoing cardiac impairment<br>n=41 | Resolved cardiac impairment<br>n=30 | P |
| **Elevated T1** | 46 (45%) | 0 (0%) | **<0.001** | 13 (68%) | 33 (40%) | **0.02** | 13 (32%) | 0 (0%) | **<0.001** |
| **Left Ventricle** |  |  |  |  |  |  |  |  |  |
| High end diastolic volume | 4 (3.9%) | 0 (0%) | **0.001** | 1 (5.3%) | 3 (3.6%) | 0.6 | 2 (4.9%) | 0 (0%) | 0.32 |
| High end systolic volume | 6 (5.9%) | 4 (0.9%) | **0.005** | 1 (5.3%) | 5 (6.0%) | 1 | 3 (7.3%) | 0 (0%) | 0.15 |
| Low ejection fraction | 21 (21%) | 0 (0%) | **<0.001** | 5 (26%) | 16 (19%) | 0.5 | 9 (22%) | 0 (0%) | **0.008** |
| High stroke volume | 1 (1.0%) | 3 (0.7%) | 0.58 | 0 (0%) | 1 (1.2%) | 1 | 0 (0%) | 0 (0%) | 1 |
| High ventricular muscle mass | 6 (5.9%) | 18 (4.2%) | 0.44 | 2 (11%) | 4 (4.8%) | 0.3 | 2 (4.9%) | 2 (6.7%) | 0.60 |
| High Ventricular maximum wall thickness | 11 (11%) | 26 (6.1%) | 0.09 | 3 (17%) | 8 (9.6%) | 0.4 | 4 (9.8%) | 4 (13%) | 0.23 |
| Low Global circumferential strain | 11 (11%) | 13 (3.1%) | **0.002** | 2 (11%) | 9 (11%) | 1 | 6 (15%) | 1 (3.3%) | 0.17 |
| Low Global longitudinal strain | 21 (21%) | 0 (0%) | **<0.001** | 1 (5.6%) | 20 (25%) | 0.1 | 7 (17%) | 0 (0%) | **0.02** |
| **Right Ventricle** |  |  |  |  |  |  |  |  |  |
| High end diastolic volume | 6 (5.9%) | 0 (0%) | **<0.001** | 2 (11%) | 4 (4.8%) | 0.3 | 3 (7.3%) | 0 (0%) | 0.15 |
| High end systolic volume | 7 (6.9%) | 2 (0.5%) | **<0.001** | 3 (16%) | 4 (4.8%) | 0.1 | 3 (7.3%) | 2 (6.7%) | 0.79 |
| Low ejection fraction | 21 (21%) | 0 (0%) | **<0.001** | 4 (21%) | 17 (20%) | 1 | 12 (29%) | 0 (0%) | **<0.001** |
| High stroke volume | 4 (3.9%) | 0 (0%) | **0.001** | 1 (5.3%) | 3 (3.6%) | 0.6 | 2 (4.9%) | 0 (0%) | 0.32 |

### From acute COVID-19 hospitalisation to 1-year trajectory

People with cardiac impairment were more likely to be male than those without cardiac impairment (41% vs 24%, p=0.001) (**Table 1**). In those with cardiac impairment, acute COVID-19 hospitalisation (19%) was associated with severe symptoms (68% vs 37%, p=0.01), wheezing (71% vs 44%, p=0.01), T1 elevation (68% vs 40%, p=0.02) and multi-organ involvement (≥3 organs; 32% vs 9.6%, p=0.02, particularly kidney impairment), compared with non-hospitalised individuals. **(Tables 1-2)**. Follow-up CMR data was available in 330/331 individuals a median 12.7 (IQR: 11.6, 14.3) months since first symptoms. Technical success of CMR and integrated assessment was 98.8% at follow-up. At 12 months, 51/330 (15%) had cardiac impairment, all symptomatic at baseline. 71/102 individuals with cardiac impairment at 6 months had follow-up data available **(Figure 1)**.

### Resolved cardiac impairment at 12 months

At 12 months cardiac impairment had resolved in 30/71 (42%). Of these individuals, 13/30 (43%) had severe Long COVID, with less symptom burden in all but 1 (median 10 and 4 symptoms at 6 and 12 months) and 5/30 (17%) were asymptomatic (**Table 1**). The most prevalent symptoms were fatigue (63%), shortness of breath (53%), chest pain (40%), myalgia (37%) and joint pain (37%), and 9 (30%) individuals had severe breathlessness. Cardiac impairment affected quality of life (mean LVD-36 score of 0.36) and 13/30 (43%) had moderate to severe problems with usual activities. 9/30 (30%) had required acute COVID-19 hospitalisation, and 3 (10%) were hospitalised between visits. Average time off work was not significantly different between resolved and ongoing impairment groups.

At 6 months, CMR in this group showed elevation in T1 (57%), low GLS (21%) and reduced LVEF (20%), with full resolution by 1 year (**Table 2**). By 12 months, 53% had fully resolved multi-organ impairment, and only 1 individual had impairment in ≥3 organs **(Table 3)**. Alongside resolution of CMR findings, elevation of NT-proBNP observed at baseline in a single patient of 41 years had resolved by 12 months. No blood investigations were predictive of cardiac recovery **(Table S2)**.

**Table 3:** Non-cardiac organ impairment in individuals at baseline and follow-up.

|  | 6 months |  |  |  |  |  |  | 12 months |  |  |
| --- | --- | --- | --- | --- | --- | --- | --- | --- | --- | --- |
|  | Overall cohort<br>n=534 | Cardiac<br>impairment<br>n=102 | No<br>Cardiac<br>impairment<br>n=424 | P | Cardiac<br>impairment<br>and<br>hospitalised,<br>n=19 | Cardiac<br>impairment not<br>hospitalised,<br>n= 83 | P | Ongoing<br>cardiac<br>impairment<br>n=41 | Resolved<br>cardiac<br>impairment<br>n=30 | P |
| **LIVER** |  |  |  |  |  |  |  |  |  |  |
| cT1 (high) | 58 (11%) | 13 (13%) | 43 (10%) | 0.48 | 3 (16%) | 10 (12%) | 0.71 | 6 (15%) | 1 (3.4%) | 0.23 |
| PDFF (high) | 119 (24%) | 25 (26%) | 92 (24%) | 0.62 | 6 (33%) | 19 (25%) | 0.55 | 9 (23%) | 5 (17%) | 0.56 |
| Volume (high) | 35 (6.6%) | 6 (5.9%) | 29 (6.9%) | 0.72 | 2 (11%) | 4 (4.8%) | 0.31 | 3 (7.3%) | 3 (10%) | 0.69 |
| **KIDNEYS** |  |  |  |  |  |  |  |  |  |  |
| Cortex T1 both kidneys (high) | 28 (5.3%) | 8 (7.8%) | 20 (4.8%) | 0.21 | 3 (16%) | 5 (6.0%) | 0.17 | 2 (4.9%) | 0 (0%) | 0.51 |
| Volume, both kidneys (high) | 18 (3.4%) | 6 (5.9%) | 12 (2.9%) | 0.14 | 3 (16%) | 3 (3.6%) | 0.08 | 2 (4.9%) | 2 (6.9%) | 1 |
| Left cortex T1 (high) | 61 (12%) | 12 (12%) | 48 (11%) | 0.92 | 5 (26%) | 7 (8.4%) | **0.04** | 4 (9.8%) | 3 (10%) | 1 |
| Right cortex T1 (high) | 46 (8.7%) | 8 (7.8%) | 38 (9.1%) | 0.70 | 3 (16%) | 5 (6.0%) | 0.17 | 5 (12%) | 1 (3.4%) | 0.40 |
| Left volume (high) | 28 (5.3%) | 7 (6.9%) | 21 (5.0%) | 0.45 | 3 (16%) | 4 (4.8%) | 0.12 | 2 (4.9%) | 2 (6.9%) | 1 |
| Right volume (high) | 38 (7.2%) | 8 (7.8%) | 30 (7.1%) | 0.81 | 4 (21%) | 4 (4.8%) | **0.04** | 4 (9.8%) | 2 (6.9%) | 1 |
| **PANCREAS** |  |  |  |  |  |  |  |  |  |  |
| T1 (high) | 46 (9.1%) | 8 (8.4%) | 37 (9.2%) | 0.81 | 2 (11%) | 6 (7.8%) | 0.64 | 2 (5.9%) | 4 (17%) | 0.22 |
| PDFF (high) | 77 (15%) | 14 (14%) | 62 (15%) | 0.86 | 4 (22%) | 10 (12%) | 0.28 | 4 (11%) | 6 (23%) | 0.30 |
| **SPLEEN** |  |  |  |  |  |  |  |  |  |  |
| Volume (high) | 42 (7.9%) | 6 (5.9%) | 36 (8.6%) | 0.37 | 1 (5.3%) | 5 (6.0%) | 1 | 4 (9.8%) | 1 (3.4%) | 0.39 |
| Length (high) | 43 (8.1%) | 8 (7.8%) | 35 (8.3%) | 0.88 | 1 (5.3%) | 7 (8.4%) | 1 | 3 (7.3%) | 0 (0%) | 0.26 |
| **LUNGS** |  |  |  |  |  |  |  |  |  |  |
| Total deep fractional area change (low) | 12 (2.4%) | 10 (2.5%) | 2 (2.2%) | 1 | 0 (0%) | 2 (2.7%) | 1 | 0 (0%) | 0 (0%) | 1 |

**Table 4:** Detailed findings of cardiac magnetic resonance at 6 and 12 months in individuals with ongoing and resolved cardiac impairment.

|  |  |  | 6 months |  |  | 12 months |  |  |
| --- | --- | --- | --- | --- | --- | --- | --- | --- |
|  |  | Healthy controls | Cardiac impairment ongoing at 12 months, n=41 | Cardiac impairment resolving at 12 months, n=41 | P | Cardiac impairment ongoing at 12 months, N=41 | Cardiac impairment resolving at 12 months, N=41 | P |
| Global T1 (ms) | 1.5T | 968 (962, 988) | 974 (35) | 987 (31) | 0.2 | 982 (26) | 970 (36) | 0.2 |
|  | 3T | 1,172 (1,150, 1,192) | 1196 (37) | 1200 (27) | 0.8 | 1200 (1172,1209) | 1194 (1188, 1204) | 0.9 |
| Left Ventricle |  |  |  |  |  |  |  |  |
| End diastolic volume (mL) |  | 86 (79, 97) | 88 (73, 97) | 80 (70, 88) | 0.18 | 86 (16) | 82 (17) | 0.30 |
| End systolic volume (mL) |  | 35 (31, 41) | 39 (30, 46) | 34 (28, 41) | 0.07 | 37 (10) | 33 (8) | 0.07 |
| Ejection fraction (%) |  | 59.5 (56.6, 62.7) | 55.0 (5.8) | 58.1 (6.0) | **0.04** | 57.7 (6.0) | 60.0 (3.9) | **0.05** |
| Stroke volume (mL) |  | 52 (46, 58) | 45 (40, 53) | 46 (43, 50) | 0.99 | 48 (43, 54) | 48 (42, 53) | 0.96 |
| Ventricular muscle mass (mm) |  | 78 (64, 96) | 87 (19) | 85 (24) | 0.72 | 86 (76, 100) | 84 (68, 108) | 0.79 |
| Ventricular max wall thickness (mm) |  | 8.91 (8.16, 10.20) | 9.45 (8.46, 10.50) | 9.24 (8.30, 10.33) | 0.50 | 9.75 (8.77, 10.74) | 9.61 (8.64, 10.58) | 0.56 |
| Global circumferential strain (%) |  | -21.28 (2.31) | -19.64 (2.67) | -21.16 (2.44) | **0.02** | -20.43 (2.68) | -21.34 (2.16) | 0.13 |
| Global longitudinal strain (%) |  | -14.68 (-15.95, -13.69) | -12.85 (-14.56, -11.49) | -13.93 (-15.03, -11.89) | 0.28 | -13.29 (2.59) | -14.49 (2.13) | **0.04** |
| Right Ventricle |  |  |  |  |  |  |  |  |
| End diastolic volume (mL) |  | 87 (78, 101) | 83 (69, 95) | 80 (67, 88) | 0.52 | 81 (72, 98) | 82 (68, 91) | 0.43 |
| End systolic volume (mL) |  | 38 (31, 45) | 37 (30, 46) | 34 (27, 39) | 0.16 | 36 (29, 43) | 34 (24, 40) | 0.18 |
| Ejection fraction (%) |  | 57.6 (4.5) | 54.9 (5.7) | 57.3 (5.5) | 0.09 | 56.1 (6.1) | 58.9 (5.1) | **0.04** |
| High stroke volume (mL) |  | 50 (45, 58) | 44 (39, 52) | 46 (39, 49) | 0.66 | 46 (40, 53) | 46 (41, 50) | 0.96 |

Abbreviations: ms, milliseconds; mm, millimetres; mL, millilitres; %, percentage. Values are presented as mean (SD) and p values calculated with T-test when the data was normally distributed. For variables where data was not normally distributed data is presented with median (IQR) and p values are calculated with Wilcoxon rank sum test.

### Ongoing and new cardiac impairment at 12 months

At 12 months cardiac impairment persisted in 58% (41/71). Symptoms and impact on usual activities and quality of life were similar with ongoing and resolved cardiac impairment at 12 months. 16/41 (39%) with ongoing impairment had severe Long COVID at 12 months, with reduced symptoms (median 10 and 2 symptoms at 6 and 12 months); 6/41 (15%) patients were asymptomatic (**Table 1**). 7/41 (17%) individuals had acute COVID-19 hospitalisation. Only 1/41 (2%) required hospitalisation between visits.

Organ impairment was more common, compared to those with resolved cardiac impairment. At 6 months, reduced LVEF (p=0.04) and low GLS (p=0.02) were more common and at 12 months, LVEF, GLS and RVEF were consistently lower (p=0.05, p=0.04 and p=0.04, respectively) (**Table 3**). One individual had abnormal T2 imaging at 12 months. Multi-organ impairment was more common (≥2 organs impaired in 49% with ongoing cardiac impairment, p=0.002); **Table 1**). Most blood investigations were within normal range and those that were abnormal (potassium, LDL, MCHC, CK, albumin, HDL, and LDL cholesterol) were also abnormal in the resolved group **(Table S2)**. Ten individuals with normal cardiac function at 6 months developed cardiac impairment by 12 months (elevated cardiac T1: n=6, low RVEF: n=4, low LVEF: n=1) **(Table S3)**.00

### Prediction

Low LVEF (p=0.03) and CRP levels ≤ 3 mg/L (p=0.019) at 6 months were significantly associated with ongoing cardiac impairment at 12 months, based on stepwise logistic regression. Cardiac impairment at 6 months did not predict any clinical outcome measures at 12 months. Low GLS and elevated cardiac T1 at 6 months were predictive of poor quality of life (OR: 0.78 (CI 0.67-0.91), p=0.001) and lower symptom severity (OR: 0.71 (CI 0.52-0.96), p=0.02) at 12 months **(Figure 2)**.

## Discussion

In the largest cohort to-date with cardiac MR follow-up over 1-year in a mainly non-hospitalised, post-COVID-19 cohort in a non-acute setting, we report three new findings. First, cardiac impairment, predominantly non-myocarditis, was common (1 in 5 individuals at 6 months) and commonly persisted (3 out of 5 individuals at 12 months). Second, cardiac impairment was found even without acute COVID hospitalisation (83/454, 18%). Third, cardiac blood biomarkers and symptoms were not predictive of cardiac impairment. Fourth, CMR parameters (e.g. left ventricular ejection fraction, 3D global longitudinal strain and cardiac T1) were predictive of quality of life and symptom severity at 12 months.

### Characteristics and trajectory of cardiac impairment

Our results indicate that, despite women being more affected by Long COVID, men have higher risk of cardiac impairment(19). Potential contributory factors include: influence of biological sex on expression and regulation of angiotensin-converting enzyme 2, sex-differences in genetic and hormonal regulation of immune responses(20), sex-dependent patterns of coagulation, smoking or drinking(11–13)(22).

Published CMR studies in Long COVID vary by study design, cohort, follow-up duration, definition of cardiac impairment, and estimated prevalence of cardiac impairment (26%-60%)(8,9,14,23–26). These have been reviewed and summarised recently (15). When COVID-19-related and classical myocardial injury are compared(27), only 9% of individuals fulfil acute myocarditis criteria and those with more severe disease are more likely to exhibit chronic inflammation and impaired cardiac function. We report prevalence of cardiac impairment (19% and 15% at 6 and 12 months) consistent with previous studies, providing standardisation of metrics and definition, which can be used at scale in research and practice to document and monitor cardiac impairment(8,9,14, 21, 24). We confirm that abnormalities in T1 (in line with previous research(8,9,14,15,25), T2 and LGE, as well as functional abnormalities (8,13,26,28) are most common in Long COVID patients. Acute Covid can present with myocardial inflammation; ongoing COVID-19 patients can also have myocarditis, but it is harder to diagnose, and often missed with echocardiography. More pertinently, the functional changes we see in this study may be due to inflammation and other aetiologies (e.g., pulmonary disease, micro-infarctions, metabolic dysregulation), and further mechanistic work is undoubtedly required.

In 58 hospitalised individuals, 3 months post-COVID-19, there were persistent abnormalities in cardiac T1(26%) and multiple organs (e.g. 29% with increased cortical T1, a marker of kidney inflammation). At 6 months, 52% had persistent symptoms and cardiac impairment(9). In the first 201 individuals from our study, we observed multi-organ impairment (29%; cardiac:26%; renal: 4%)(8). In 443 individuals 10 months after mild-to-moderate COVID-19, subclinical multi-organ impairment was associated with cardiac impairment (reduced left and right ventricular systolic function)(10). At 12 months, the longest follow-up duration to-date, we now show that 54% of individuals with cardiac impairment do not fully recover.

### Impact of acute hospitalisation for COVID-19 on cardiac impairment

Most individuals presenting with cardiac injury at baseline in this trial did not require acute COVID-19 hospitalization (81%). Only 1 patient with elevated cardiac-related blood biomarkers had CMR abnormalities at 6 months and acute COVID-19 hospitalisation. Blood biomarkers and symptoms did not differentiate hospitalised and non-hospitalized groups. On MRI assessment, cardiac T1 abnormalities(12,29) and multi-organ involvement (particularly renal)(8,9,13) were more prevalent in those with cardiac impairment and acute COVID-19 hospitalisation, as in other published studies.

### Clinical management pathways in Long COVID populations at risk of cardiac impairment

Cardiac-related blood biomarkers may be raised in early convalescence from COVID-19(30), but did not aid detection of cardiac impairment in Long COVID in our study, despite 19% having CMR abnormalities, supported by other research(13,26,28). Burden and improvement in symptoms 6 months after COVID-19 were correlated with neither improvement on CMR nor lung parenchymal recovery (13). There are implications for roles of blood investigations and imaging in diagnosis and follow up of cardiac and multi-organ impairment in Long COVID, to minimise burden to healthcare systems, already struggling due to COVID-19-related lack of resources and backlogs, while achieving integrated care.

A rapid and complete multi-organ assessment such as COVERSCAN may help clinical decision-making and improve healthcare access and provision. For example, a negative COVERSCAN could rule out need for cardiology referral, even in presence of symptoms (e.g. chest pain, shortness of breath). A positive COVERSCAN could guide follow up assessment and identification of Long COVID subtypes in research and practice. Interventional trials with pre-specified subgroup analysis and improved definitions of cardiac impairment (not only myocarditis-centred), are required to inform cost-effective therapies.

### Strengths and Limitations

This is the largest longitudinal study to-date of cardiac impairment in Long COVID with detailed biochemical and imaging characterisation of multi-organ function starting in April 2020. We included healthy, age-matched controls. All MRI was non-contrast. We recruited a real-world cohort at lower risk of COVID-19 severity and mortality. Unlike other studies(10), our approach offers quick, scalable assessment using standard MRI scanners. There are limitations. First, our CMR protocol excluded gadolinium contrast due to concerns regarding COVID-19-related renal complications, relying on native non-invasive T1 mapping to characterise myocardial inflammation, validated for acute myocarditis(31). Second, we did not have follow-up scans on individuals without impairment at baseline. Third, we did not have pre-COVID cardiac or multi-organ imaging available in participants. Third, our study population was not ethnically diverse, and COVID-19 has disproportionately affected non-white individuals.

### Conclusion

CMR shows that cardiac impairment persists in Long Covid in some individuals up to 12 months after first symptoms. Cardiac impairment is associated with acute COVID-19 hospitalisation and male gender, but subtypes of disease (based on symptoms, examination, and investigations) are yet to be established. Therapeutic options and effective clinical pathways require urgent clinical trials.

## Data Availability

All data produced in the present work are contained in the manuscript

## Acknowledgments

The study was partly financed by an Amendment to a European Commission’s Horizon 2020 grant 719445 (AMENDMENT Reference No AMD-719445-8): Non-invasive rapid assessment of chronic liver disease using Magnetic Resonance Imaging with LiverMultiScan (RADIcAL). AB has received funding from NIHR (including the STIMULATE-ICP study), AstraZeneca, European Union, UK Research and Innovation and British Medical Association. Conflicts of interest: AD, AT, VC, ABo, SF, MP, ARF, HTB, MK, MR, MB and RB are employees of Perspectum.

## Ethical Approval

The protocol received full ethical approval from South Central - Berkshire B Research Ethics Committee (20/SC/0185) and was registered (https://clinicaltrials.gov/ct2/show/NCT04369807).

## Data availability

All data produced in the present work are contained in the manuscript.

