## Supplementary methods, tables and figures for "Cardiac impairment in Long Covid 1-year post-SARS-CoV-2 infection"

**Supplementary materials**

| **Page Number** | **Contents** |
| --- | --- |
| 2 | Supplementary methods |
| 5 | Table S1: Reference Ranges for imaging metrics across organs |
| 8 | Table S2: Blood investigations by cardiac impairment on MRI |
| 16 | Table S3. Detailed CMR findings in new onset cardiac impairment at 12 months. |
| 17 | Supplementary References: |

**Supplementary methods**

1. **Classification of Severity:**
   a. **Definition Long COVID, based on self-reported questionnaires** (1–3)**:**

|  | **Dyspnoea 12 score (1)** |  | **EQ-5D-5L usual activity score*(4)** |
| --- | --- | --- | --- |
| **Severe Long COVID** | ≥10 | or | ≥3 |
| **Mild Long COVID** | Not fulfilling neither the “severely symptomatic” or “asymptomatic” conditions | | |
| **Asymptomatic** | 0 | and | 0 |

*** EQ-5D-5L score is based on the scoring in the following question:**

*“Please tick the ONE box that best describes your health TODAY”*

*USUAL ACTIVITIES (e.g., work, study, housework, family, or leisure activities)*

- *I have no problems doing my usual activities (score 1)*
- *I have slight problems doing my usual activities (score 2)*
- *I have moderate problems doing my usual activities (score 3)*
- *I have severe problems doing my usual activities (score 4)*
- *I am unable to do my usual activities (score 5)*
  b. **Definition of symptom severity, based on self-reported perceived severity:**

**Patient asked to select one of the following options:**

Critical acute respiratory distress syndrome (ARDS)

Severe disease

Moderate disease

Mild disease

Asymptomatic disease

c. **Severe breathlessness**

Score of ≥10 in the Dyspnoea-12 validated questionnaire

2. **List of blood biomarkers assessed in this study:**

Haemoglobin, HCT, red cell count, MCV, MCH , MCHC, RDW , platelet count, MPV, white cell count , neutrophils, lymphocytes monocytes, eosinophils, basophils, ESR, sodium, potassium, chloride, bicarbonate, urea, creatinine, bilirubin, alkaline phosphatase, aspartate transferase , alanine transferase , LDH , CK , gamma , total protein , albumin , globulin, calcium, magnesium, phosphate, uric acid, triglycerides, fasting triglycerides, cholesterol, fasting cholesterol, HDL cholesterol, LDL cholesterol, iron, TIBC, transferrin saturation, CRP high sensitivity, troponin I high sensitivity, amylase, ferritin, lipase, thyroid stimulating hormone, testosterone, insulin , C-peptide, NTpro-BNP.

3. **Imaging acquisitions:**

- Cardiac imaging involved a combination of several gated cine series, two long axis cines (horizontal long axis – HLA and vertical long axis – VLA), and a complete short axis stack covering the left ventricle (LV) and right ventricle (RV). This acquisition mirrors the one used at the UK Biobank and is a standardized approach (5). Three short-axis were acquired at the basal, mid, and apical levels of the left ventricle, for T1 mapping using MOLLI, and for T2 mapping using a T2 preparation pulse applied with different T2 preparation times, to impart T2 signal contrast, and a subsequent readout is performed by using a steady-state free precession (SSFP) or a fast low angle shot (FLASH).
- Liver and pancreas imaging used the LiverMultiScan acquisition protocol (Perspectum, Oxford, UK), which involves 3 single 2D axial slice breath-held acquisitions that separately are sensitive to the fat content (proton density fat fraction, or PDFF), to T2* (which is representative of liver iron content) and a MOLLI-T1 measurement (providing a measurement of tissue water), additionally a volumetric scan was used that covers the entire liver (6).
- Lungs: Two dynamic cine MR acquisitions were acquired in the coronal plane with a 306.91ms temporal resolution: one 40 s acquisition with the patient instructed to breathe normally and a second 30 s acquisition with the patient instructed to breathe deeply.
- Kidney: single coronal view that was able to image both kidneys. Imaging contrasts were MOLLI-T1, and a spoiled gradient recalled acquisition (SPGR).
- Spleen: Volumetric SPGR MRI images

4. **Image Analysis:**

- Cardiac: Experienced cardiac MRI analysts used CVI42v5.11 (Cardiovascular Imaging Inc, Canada) to trace manually the myocardium in the end-diastolic and end-systolic phases in each of the short-axis views, following the standard UK Biobank evaluation approach as previously described (7). We reported ventricular function; end systolic and diastolic volume; stroke volume and ejection fraction in both ventricles; left ventricular muscle mass and ventricular max wall thickness and global longitudinal and circumferential 3D strain metrics. Mean Cardiac T1 and T2 were determined for each of the 16 cardiac segments (of the AHA 17 segment model excluding the apex)(8).
- Liver Images were analysed by data analysts experienced at using the LiverMultiScan (Perspectum, Oxford, UK) software. This yielded global metrics in each liver of PDFF (proton density fat fraction), T2*, and cT1 (cT1 is a measurement of T1 that has been corrected for the confounding effects of iron and standardised to 3 Tesla; it is elevated with disease).
- Pancreas images were analysed in an equivalent manner to the above except the software used was not FDA-cleared and iron correction was not performed. The output T1 was standardized to 3 Tesla.
- Lung cine imaging allowed the measurement of the area of the left and right lungs through the breathing cycle in the coronal plane, which used automated methods that were reviewed by image analysts. The periodicity of the area fluctuations was used to determine the respiratory rate. All analysis was performed in-house using MATLAB based tools. The method was validated by measuring the correlation between the change in area and the forced vital capacity, the latter being measured using spirometry. Patient respiration was assessed by imaging a single 2D coronal slice of the lungs over 30 seconds using a dynamic cine MRI acquisition, during which the patient instructed to breathe deeply.
- Kidney: assessed using in-house tools to fit parametric maps and to allow trained analysts to make measurements. The kidney cortex was manually segmented using the MOLLI-T1 map to guide the boundary. Multiple regions-of-interests were manually placed within the cortex to extract a median value of cortical T1 in each kidney. Volumetric delineations of the kidneys were derived from SPGR MRI images. Automated delineations were produced using a 3D convolutional neural network, trained on expert annotations. Delineations were manually checked, and corrected, if necessary, for each subject. In addition to kidney cortex T1 and kidney volume, we also derived kidney length measurements, in the inferior-superior axis, from the same organ segmentations and assessed the correlation of kidney length and kidney volume measurements
- Spleen: Volumetric delineations were derived from SPGR MRI images. Automated delineations were produced using a 3D convolutional neural network, trained on expert annotations. Delineations were manually checked, and corrected, if necessary, for each subject.
- Organ impairment: Calculated for each organ based on evidence of any of the measurements appearing out of reference range (*Liver:* elevated cT1 or Fat; *Kidney:* elevated T1 or volume; *Pancreas:* elevated sT1 or Fat; *Heart:* elevated T1 in 3 or more segments, decreased RV or LV EF or increased LV or RV EDV or increased LV global longitudinal strain; *Spleen*: elevated volume; *Lung:* reduced fractional area volume). Single organ impairment was based on ≥1 organ impairment and multi-organ impairment was based on ≥2 organ impairments.

5. **Reference Ranges for imaging markers:**

All values but organ volumes were calculated with n=92 HC (Healthy Controls) scanned at 1.5T and 3T for this study calculating 2.5% (lower threshold) and 97.5% percentiles (upper threshold). Organ volumes were calculated from a combined cohort of the 92 healthy controls and 1744 BMI matched participants (N=1836 from N=36) from the UK Biobank, (9), representing all sex and height subgroups, as these are known confounders of organ size.(10) (*) Reference ranges for the liver cT1 and liver PDFF have been taken from the available literature (11), as the LMS technology have been widely used and tested in multiple clinical trials and research settings. For pancreas PDFF, which has a positive skew in the distribution, reference ranges were extracted with the 95% percentile. ($) T2 repeatability coefficients are not provided as this metric was only available for follow up in patients. (§) Right and left cortical T1 limits were averaged for analysis (**Table S1**).

**Table S1: Reference Ranges for imaging metrics across organs**

|  | **Gender** | **Field Strength** | **Height (cm)** | **Lower threshold** | **Upper threshold (*)** | **Repeatability coefficient** |
| --- | --- | --- | --- | --- | --- | --- |
| **CARDIAC METRICS** | | | | | | |
| **Field strength independent variables (BSA corrected)** | | | | | | |
| Left end diastolic volume (mL)  Left end diastolic volume (mL)  Right end diastolic volume (mL)  Right end diastolic volume (mL)  Left end Systolic volume (mL)  Left end Systolic volume (mL)  Right end Systolic volume (mL)  Right end Systolic volume (mL)  Left Stroke volume (mL)  Left Stroke volume (mL)  Right Stroke volume (mL)  Right Stroke volume (mL) | F  M  F  M  F  M  F  M  F  M  F  M | -  -  -  -  -  -  -  -  -  -  -  - | -  -  -  -  -  -  -  -  -  -  -  - | -  -  -  -  -  -  -  -  -  -  -  - | 108  132  110  139  47  57  49  60  66  84  65  84 | 17  17  19  19  12.5  12.5  12  12  16  16  16  16 |
| **Field strength independent variables (non-BSA corrected)** | | | | | | |
| Global circumferential strain 3D (%)  Global circumferential strain 3D (%)  Global longitudinal strain 3D (%)  Global longitudinal strain 3D (%)  Left ventricle ejection fraction (%)  Left ventricle ejection fraction (%)  Right ventricle ejection fraction (%)  Right ventricle ejection fraction (%)  Left ventricular max wall thickness (mm)  Left ventricular max wall thickness (mm)  Left ventricular muscle mass (g)  Left ventricular muscle mass (g) | F  M  F  M  F  M  F  M  F  M  F  M | -  -  -  -  -  -  -  -  -  -  -  - | -  -  -  -  -  -  -  -  -  -  -  - | -  -  -  -  52  51  50  50  -  -  -  - | -18.1  -16.8  -11.5  -7.8  -  -  -  -  10.6  14  95  151 | 2.5  2.5  5.1  5.1  6.6  6.6  7.0  7.0  2.1  2.1  13  13 |
| **Field strength Dependent variables: 1.5T** | | | | | | |
| Global T1 ref range (ms)  Global T1 ref range (ms)  Segment 1: T1 basal anterior (ms)  Segment 1: T1 basal anterior (ms)  Segment 2: T1 basal anteroseptal (ms)  Segment 2: T1 basal anteroseptal (ms)  Segment 3: T1 basal inferoseptal (ms)  Segment 3: T1 basal inferoseptal (ms)  Segment 4: T1 basal inferior (ms)  Segment 4: T1 basal inferior (ms)  Segment 5: T1 basal inferolateral (ms)  Segment 5: T1 basal inferolateral (ms)  Segment 6: T1 basal anterolateral (ms)  Segment 6: T1 basal anterolateral (ms)  Segment 7: T1 mid anterior (ms)  Segment 7: T1 mid anterior (ms)  Segment 8: T1 mid anteroseptal (ms)  Segment 8: T1 mid anteroseptal (ms)  Segment 9: T1 mid inferoseptal (ms)  Segment 9: T1 mid inferoseptal (ms)  Segment 10: T1 mid inferior (ms)  Segment 10: T1 mid inferior (ms)  Segment 11: T1 mid inferolateral (ms)  Segment 11: T1 mid inferolateral (ms)  Segment 12: T1 mid anterolateral (ms)  Segment 12: T1 mid anterolateral (ms)  Segment 13: T1 apical anterior (ms)  Segment 13: T1 apical anterior (ms)  Segment 14: T1 apical septal (ms)  Segment 14: T1 apical septal (ms)  Segment 15: T1 apical inferior (ms)  Segment 15: T1 apical inferior (ms)  Segment 16: T1 apical lateral (ms)  Segment 16: T1 apical lateral (ms)  Global T2 ref range (ms) ($) | F  M  F  M  F  M  F  M  F  M  F  M  F  M  F  M  F  M  F  M  F  M  F  M  F  M  F  M  F  M  F  M  F  M  - | 1.5T  1.5T  1.5T  1.5T  1.5T  1.5T  1.5T  1.5T  1.5T  1.5T  1.5T  1.5T  1.5T  1.5T  1.5T  1.5T  1.5T  1.5T  1.5T  1.5T  1.5T  1.5T  1.5T  1.5T  1.5T  1.5T  1.5T  1.5T  1.5T  1.5T  1.5T  1.5T  1.5T  1.5T  1.5T | -  -  -  -  -  -  -  -  -  -  -  -  -  -  -  -  -  -  -  -  -  -  -  -  -  -  -  -  -  -  -  -  -  -  - | -  -  -  -  -  -  -  -  -  -  -  -  -  -  -  -  -  -  -  -  -  -  -  -  -  -  -  -  -  -  -  -  -  -  - | 1042  997  1043  1000  1031  1022  1031  1001  1091  995  1042  998  1041  979  1014  969  1030  1006  1036  994  1035  1023  1016  982  1029  979  1059  1004  1065  992  1070  1003  1040  1011  51 | -  -  42  42  9  49  54  54  57  57  55  55  54  54  52  52  39  39  37  37  44  44  44  44  62  62  86  86  48  48  43  43  70  70  - |
| **Field strength Dependent variables: 3T** | | | | | | |
| Global T1 ref range (ms)  Global T1 ref range (ms)  Segment 1: T1 basal anterior (ms)  Segment 1: T1 basal anterior (ms)  Segment 2: T1 basal anteroseptal (ms)  Segment 2: T1 basal anteroseptal (ms)  Segment 3: T1 basal inferoseptal (ms)  Segment 3: T1 basal inferoseptal (ms)  Segment 4: T1 basal inferior (ms)  Segment 4: T1 basal inferior (ms)  Segment 5: T1 basal inferolateral (ms)  Segment 5: T1 basal inferolateral (ms)  Segment 6: T1 basal anterolateral (ms)  Segment 6: T1 basal anterolateral (ms)  Segment 7: T1 mid anterior (ms)  Segment 7: T1 mid anterior (ms)  Segment 8: T1 mid anteroseptal (ms)  Segment 8: T1 mid anteroseptal (ms)  Segment 9: T1 mid inferoseptal (ms)  Segment 9: T1 mid inferoseptal (ms)  Segment 10: T1 mid inferior (ms)  Segment 10: T1 mid inferior (ms)  Segment 11: T1 mid inferolateral (ms)  Segment 11: T1 mid inferolateral (ms)  Segment 12: T1 mid anterolateral (ms)  Segment 12: T1 mid anterolateral (ms)  Segment 13: T1 apical anterior (ms)  Segment 13: T1 apical anterior (ms)  Segment 14: T1 apical septal (ms)  Segment 14: T1 apical septal (ms)  Segment 15: T1 apical inferior (ms)  Segment 15: T1 apical inferior (ms)  Segment 16: T1 apical lateral (ms)  Segment 16: T1 apical lateral (ms)  Global T2 ref range (ms) ($) | F  M  F  M  F  M  F  M  F  M  F  M  F  M  F  M  F  M  F  M  F  M  F  M  F  M  F  M  F  M  F  M  F  M  - | 3T  3T  3T  3T  3T  3T  3T  3T  3T  3T  3T  3T  3T  3T  3T  3T  3T  3T  3T  3T  3T  3T  3T  3T  3T  3T  3T  3T  3T  3T  3T  3T  3T  3T  3T | -  -  -  -  -  -  -  -  -  -  -  -  -  -  -  -  -  -  -  -  -  -  -  -  -  -  -  -  -  -  -  -  -  -  - | -  -  -  -  -  -  -  -  -  -  -  -  -  -  -  -  -  -  -  -  -  -  -  -  -  -  -  -  -  -  -  -  -  -  - | 1255  1214  1226  1201  1248  1218  1251  1218  1271  1231  1240  1209  1200  1193  1266  1161  1264  1219  1272  1226  1279  1228  1226  1210  1278  1228  1271  1227  1280  1230  1257  1202  1254  1214  46 | -  -  72  72  70  70  74  74  112  112  109  109  61  61  90  90  89  89  74  74  84  84  60  60  75  75  63  63  62  62  57  57  77  77  - |
| **LIVER METRICS** | | | | | | |
| **Field strength independent variables** | | | | | | |
| cT1 ROI (ms)  PDFF %  Volume (mL)  Volume (mL)  Volume (mL)  Volume (mL) | -  -  F  M  F  M | -  -  -  -  -  - | -  -  <164  <164  ≥ 164, < 250  ≥ 164, < 250 | -  -  -  -  -  - | 800 (*)  5 (*)  1778  2003  2049  2284 | 48  1.5  64  64  64  64 |
| **KIDNEY METRICS** | | | | | | |
| **Field strength independent variables** | | | | | | |
| Left Volume (mL)  Left Volume (mL)  Left Volume (mL)  Left Volume (mL)  Right Volume (mL)  Right Volume (mL)  Right Volume (mL)  Right Volume (mL) | F  M  F  M  F  M  F  M | -  -  -  -  -  -  -  - | <164  <164  ≥ 164, < 250  ≥ 164, < 250  <164  <164  ≥ 164, < 250  ≥ 164, < 250 | -  -  -  -  -  -  -  - | 177  221  192.  255  176  207  186  229 | 10  10  10  10  8  8  8  8 |
| **Field strength Dependent variables: 1.5T** | | | | | | |
| Cortex T1 (ms) (§) | - | 1.5T | - | - | 1154 | 76 |
| **Field strength Dependent variables: 3T** | | | | | | |
| Cortex T1 (ms) (§) | - | 3T | - | - | 1512 | 68 |
| **PANCREAS** | | | | | | |
| **Field strength independent variables** | | | | | | |
| sT1 ROI (ms)  PDFF % | -  - | -  - | -  - | -  - | 821  6.6 (*) | 74  2.8 |
| **SPLEEN** | | | | | | |
| **Field strength independent variables** | | | | | | |
| Volume (mL)  Volume (mL)  Volume (mL)  Volume (mL) | F  M  F  M | -  -  -  - | <164  <164  ≥ 164, < 250  ≥ 164, < 250 | -  -  -  - | 254  392  293  411 | 17  17  17  17. |
| **LUNG** | | | | | | |
| **Field strength independent variables** | | | | | | |
| Total deep fractional area change (%) | - | - | - | 22 | - | 15.9 |

**Table S2: Blood investigations by cardiac impairment on MRI**

|  | **6 months** | | | | | | | | | | **12 months** | | |
| --- | --- | --- | --- | --- | --- | --- | --- | --- | --- | --- | --- | --- | --- |
|  | **COVID,**  **N=534** | **No Cardiac Impairment,**  **N = 424** | **Cardiac Impairment,**  **N = 102** | **P** | **Cardiac Injury hospitalised,**  **N = 19** | **Cardiac Injury non hospitalized,**  **N = 83** | **P** | **Ongoing Cardiac injury,**  **N = 41** | **Resolved Cardiac injury,**  **N = 30** | **P** | **Ongoing Cardiac injury,**  **N = 41** | **Resolved Cardiac injury,**  **N = 30** | **P** |
| **Haemoglobin** | | | | | | | | | | | | | |
| H | 6 (1.2%) | 4 (1.0%) | 2 (2.2%) | 0.36 | 0 (0%) | 2 (2.6%) | 0.60 | 1 (2.6%) | 0 (0%) | 0.51 | 1 (2.4%) | 0 (0%) | 1.00 |
| L | 13 (2.6%) | 9 (2.2%) | 3 (3.3%) |  | 1 (6.7%) | 2 (2.6%) |  | 2 (5.3%) | 0 (0%) |  | 1 (2.4%) | 0 (0%) |  |
| N | 487 (96%) | 393 (97%) | 87 (95%) |  | 14 (93%) | 73 (95%) |  | 35 (92%) | 27 (100%) |  | 39 (95%) | 27 (100%) |  |
| Missing | 28 | 18 | 10 |  | 4 | 6 |  | 3 | 3 |  | 0 | 3 |  |
| **HCT** | | | | | | | | | | | | | |
| H | 10 (2.0%) | 9 (2.2%) | 1 (1.1%) | 0.69 | 0 (0%) | 1 (1.3%) | 0.42 | 1 (2.6%) | 0 (0%) | 1.00 | 1 (2.4%) | 2 (7.4%) | 0.56 |
| L | 8 (1.6%) | 5 (1.2%) | 2 (2.2%) |  | 1 (6.7%) | 1 (1.3%) |  | 1 (2.6%) | 0 (0%) |  | 0 (0%) | 0 (0%) |  |
| N | 488 (96%) | 392 (97%) | 89 (97%) |  | 14 (93%) | 75 (97%) |  | 36 (95%) | 27 (100%) |  | 40 (98%) | 25 (93%) |  |
| Missing | 28 | 18 | 10 |  | 4 | 6 |  | 3 | 3 |  | 0 | 3 |  |
| **Red cell count** | | | | | | | | | | | | | |
| H | 14 (2.8%) | 13 (3.2%) | 1 (1.1%) | 0.69 | 0 (0%) | 1 (1.3%) | 0.52 | 0 (0%) | 0 (0%) | 0.51 | 1 (2.4%) | 1 (3.7%) | 0.38 |
| L | 17 (3.4%) | 12 (3.0%) | 3 (3.3%) |  | 1 (6.7%) | 2 (2.6%) |  | 2 (5.3%) | 0 (0%) |  | 3 (7.3%) | 0 (0%) |  |
| N | 475 (94%) | 381 (94%) | 88 (96%) |  | 14 (93%) | 74 (96%) |  | 36 (95%) | 27 (100%) |  | 37 (90%) | 26 (96%) |  |
| Missing | 28 | 18 | 10 |  | 4 | 6 |  | 3 | 3 |  | 0 | 3 |  |
| **MCV** | | | | | | | | | | | | | |
| H | 1 (0.2%) | 0 (0%) | 1 (1.1%) | 0.20 | 0 (0%) | 1 (1.3%) | 1.00 | 1 (2.6%) | 0 (0%) | 0.51 | 2 (4.9%) | 0 (0%) | 0.30 |
| L | 10 (2.0%) | 8 (2.0%) | 2 (2.2%) |  | 0 (0%) | 2 (2.6%) |  | 2 (5.3%) | 0 (0%) |  | 0 (0%) | 1 (3.7%) |  |
| N | 495 (98%) | 398 (98%) | 89 (97%) |  | 15 (100%) | 74 (96%) |  | 35 (92%) | 27 (100%) |  | 39 (95%) | 26 (96%) |  |
| Missing | 28 | 18 | 10 |  | 4 | 6 |  | 3 | 3 |  | 0 | 3 |  |
| **MCH** | | | | | | | | | | | | | |
| H | 4 (0.8%) | 3 (0.7%) | 1 (1.1%) | 0.84 | 0 (0%) | 1 (1.3%) | 1.00 | 1 (2.6%) | 0 (0%) | 1.00 | 1 (2.4%) | 0 (0%) | 1.00 |
| L | 8 (1.6%) | 7 (1.7%) | 1 (1.1%) |  | 0 (0%) | 1 (1.3%) |  | 1 (2.6%) | 0 (0%) |  | 1 (2.4%) | 1 (3.7%) |  |
| N | 494 (98%) | 396 (98%) | 90 (98%) |  | 15 (100%) | 75 (97%) |  | 36 (95%) | 27 (100%) |  | 39 (95%) | 26 (96%) |  |
| Missing | 28 | 18 | 10 |  | 4 | 6 |  | 3 | 3 |  | 0 | 3 |  |
| **MCHC** | | | | | | | | | | | | | |
| H | 105 (21%) | 76 (19%) | 26 (28%) | **0.04** | 2 (13%) | 24 (31%) | 0.22 | 13 (34%) | 4 (15%) | 0.08 | 10 (24%) | 2 (7.4%) | 0.07 |
| L | 0 (0%) | 0 (0%) | 0(0%) |  | 0 (0%) | 0 (0%) |  | 0 (0%) | 0 (0%) |  | 0 (0%) | 0 (0%) |  |
| N | 401 (79%) | 330 (81%) | 66 (72%) |  | 13 (87%) | 53 (69%) |  | 25 (66%) | 23 (85%) |  | 31 (76%) | 25 (93%) |  |
| Missing | 28 | 18 | 10 |  | 4 | 6 |  | 3 | 3 |  | 0 | 3 |  |
| **RDW** | | | | | | | | | | | | | |
| H | 12 (2.4%) | 10 (2.5%) | 2 (2.2%) | 0.94 | 0 (0%) | 2 (2.6%) | 1.00 | 2 (5.3%) | 0 (0%) | 0.53 | 0 (0%) | 0 (0%) | 1.00 |
| L | 26 (5.1%) | 22 (5.4%) | 4 (4.3%) |  | 0 (0%) | 4 (5.2%) |  | 3 (7.9%) | 1 (3.7%) |  | 1 (2.4%) | 0 (0%) |  |
| N | 467 (92%) | 373 (92%) | 86 (93%) |  | 15 (100%) | 71 (92%) |  | 33 (87%) | 26 (96%) |  | 40 (98%) | 27 (100%) |  |
| Missing | 29 | 19 | 10 |  | 4 | 6 |  | 3 | 3 |  | 0 | 3 |  |
| **Platelet count** | | | | | | | | | | | | | |
| H | 22 (4.4%) | 15 (3.7%) | 6 (6.6%) | 0.13 | 1 (6.7%) | 5 (6.6%) | 1.00 | 4 (11%) | 1 (3.7%) | 0.49 | 1 (2.4%) | 3 (11%) | 0.29 |
| L | 2 (0.4%) | 1 (0.2%) | 1 (1.1%) |  | 0(0%) | 1 (1.3%) |  | 1 (2.7%) | 0(0%) |  | 0(0%) | 0(0%) |  |
| N | 479 (95%) | 388 (96%) | 84 (92%) |  | 14 (93%) | 70 (92%) |  | 32 (86%) | 26 (96%) |  | 40 (98%) | 24 (89%) |  |
| Missing | 31 | 20 | 11 |  | 4 | 7 |  | 4 | 3 |  | 0 | 3 |  |
| **MPV** | | | | | | | | | | | | | |
| H | 8 (1.6%) | 6 (1.5%) | 1 (1.1%) | 1.00 | 0 (0%) | 1 (1.3%) | 1.00 | 1 (2.6%) | 0  (0%) | 1.00 | 2 (4.9%) | 0 (0%) | 0.51 |
| L | 0 (0%) | 0 (0%) | 0 (0%) |  | 0 (0%) | 0 (0%) |  | 0(0%) | 0 (0%) |  | 0 (0%) | 0 (0%) |  |
| N | 496 (98%) | 398 (99%) | 91 (99%) |  | 15 (100%) | 76 (99%) |  | 37 (97%) | 27 (100%) |  | 39 (95%) | 27 (100%) |  |
| Missing | 30 | 20 | 10 |  | 4 | 6 |  | 3 | 3 |  | 0 | 3 |  |
| **White cell count** | | | | | | | | | | | | | |
| H | 19 (3.8%) | 13 (3.2%) | 3 (3.3%) | 1.00 | 0 (0%) | 3 (3.9%) | 1.00 | 2 (5.3%) | 0 (0%) | 0.51 | 0 (0%) | 0 (0%) | 1.00 |
| L | 1 (0.2%) | 1 (0.2%) | 0 (0%) |  | 0 (0%) | 0 (0%) |  | 0 (0%) | 0 (0%) |  | 0 (0%) | 0 (0%) |  |
| N | 486 (96%) | 392 (97%) | 89 (97%) |  | 15 (100%) | 74 (96%) |  | 36 (95%) | 27 (100%) |  | 41 (100%) | 27 (100%) |  |
| Missing | 28 | 18 | 10 |  | 4 | 6 |  | 3 | 3 |  | 0 | 3 |  |
| **Neutrophils** | | | | | | | | | | | | | |
| H | 8 (1.6%) | 7 (1.7%) | 1 (1.1%) | 0.60 | 0 (0%) | 1 (1.3%) | 0.66 | 1 (2.6%) | 0 (0%) | 0.78 | 0 (0%) | 0 (0%) | 1.00 |
| L | 30 (5.9%) | 20 (4.9%) | 7 (7.6%) |  | 0 (0%) | 7 (9.1%) |  | 3 (7.9%) | 1 (3.7%) |  | 3 (7.3%) | 2 (7.4%) |  |
| N | 468 (92%) | 379 (93%) | 84 (91%) |  | 15 (100%) | 69 (90%) |  | 34 (89%) | 26 (96%) |  | 38 (93%) | 25 (93%) |  |
| Missing | 28 | 18 | 10 |  | 4 | 6 |  | 3 | 3 |  | 0 | 3 |  |
| **Lymphocytes** | | | | | | | | | | | | | |
| H | 2 (0.4%) | 1 (0.2%) | 1 (1.1%) | 0.27 | 0 (0%) | 1 (1.3%) | 0.65 | 0 (0%) | 1 (3.7%) | 0.59 | 1 (2.4%) | 1 (3.7%) | 0.22 |
| L | 38 (7.5%) | 32 (7.9%) | 5 (5.4%) |  | 0 (0%) | 5 (6.5%) |  | 2 (5.3%) | 2 (7.4%) |  | 7 (17%) | 1 (3.7%) |  |
| N | 466 (92%) | 373 (92%) | 86 (93%) |  | 15 (100%) | 71 (92%) |  | 36 (95%) | 24 (89%) |  | 33 (80%) | 25 (93%) |  |
| Missing | 28 | 18 | 10 |  | 4 | 6 |  | 3 | 3 |  | 0 | 3 |  |
| **Monocytes** | | | | | | | | | | | | | |
| H | 4 (0.8%) | 2 (0.5%) | 1 (1.1%) | 0.64 | 0 (0%) | 1 (1.3%) | 1.00 | 1 (2.6%) | 0 (0%) | 1.00 | 0 (0%) | 0 (0%) | 1.00 |
| L | 2 (0.4%) | 2 (0.5%) | 0 (0%) |  | 0 (0%) | 0 (0%) |  | 0 (0%) | 0 (0%) |  | 0 (0%) | 0 (0%) |  |
| N | 500 (99%) | 402 (99%) | 91 (99%) |  | 15 (100%) | 76 (99%) |  | 37 (97%) | 27 (100%) |  | 41 (100%) | 27 (100%) |  |
| Missing | 28 | 18 | 10 |  | 4 | 6 |  | 3 | 3 |  | 0 | 3 |  |
| **Eosinophils** | | | | | | | | | | | | | |
| H | 14 (2.8%) | 10 (2.5%) | 4 (4.3%) | 0.30 | 0 (0%) | 4 (5.2%) | 1.00 | 0 (0%) | 1 (3.7%) | 0.42 | 2 (4.9%) | 1 (3.7%) | 1.00 |
| L | 0 (0%) | 0 (0%) | 0 (0%) |  | 0 (0%) | 0 (0%) |  | 0 (0%) | 0 (0%) |  | 0 (0%) | 0 (0%) |  |
| N | 492 (97%) | 396 (98%) | 88 (96%) |  | 15 (100%) | 73 (95%) |  | 38 (100%) | 26 (96%) |  | 39 (95%) | 26 (96%) |  |
| Missing | 28 | 18 | 10 |  | 4 | 6 |  | 3 | 3 |  | 0 | 3 |  |
| **Basophils** | | | | | | | | | | | | | |
| H | 2 (0.4%) | 2 (0.5%) | 0 (0%) | 1.00 | 0 (0%) | 0 (0%) | 1.0 | 0 (0%) | 0 (0%) | 1.00 | 0 (0%) | 2 (7.4%) | 0.15 |
| L | 0 (0%) | 0(0%) | 0 (0%) |  | 0 (0%) | 0 (0%) |  | 0 (0%) | 0 (0%) |  | 0 (0%) | 0 (0%) |  |
| N | 504(100%) | 404 (100%) | 92 (100%) |  | 15 (100%) | 77 (100%) |  | 38 (100%) | 27 (100%) |  | 41 (100%) | 25 (93%) |  |
| Missing | 28 | 18 | 10 |  | 4 | 6 |  | 3 | 3 |  | 0 | 3 |  |
| **ESR** | | | | | | | | | | | | | |
| H | 40 (7.9%) | 32 (7.9%) | 8 (8.6%) | 0.81 | 1 (6.7%) | 7 (9.0%) | 1.00 | 4 (11%) | 3 (11%) | 1.00 | 3 (7.3%) | 4 (15%) | 1.0 |
| L | 0 (0%) | 0 (0%) | 0 (0%) |  | 0 (0%) | 0 (0%) |  | 0 (0%) | 0 (0%) |  | 0 (0%) | 0 (0%) |  |
| N | 468 (92%) | 375 (92%) | 85 (91%) |  | 14 (93%) | 71 (91%) |  | 34 (89%) | 24 (89%) |  | 38 (93%) | 23 (85%) |  |
| Missing | 26 | 17 | 9 |  | 4 | 5 |  | 3 | 3 |  | 0 | 3 |  |
| **Sodium** | | | | | | | | | | | | | |
| H | 1 (0.2%) | 1 (0.2%) | 0 (0%) | 0.62 | 0 (0%) | 0 (0%) | 1.00 | 0 (0%) | 0 (0%) | 0.13 | 0 (0%) | 0 (0%) | 0.14 |
| L | 17 (3.4%) | 13 (3.2%) | 4 (4.3%) |  | 0 (0%) | 4 (5.2%) |  | 4 (11%) | 0 (0%) |  | 4 (9.8%) | 0 (0%) |  |
| N | 488 (96%) | 392 (97%) | 88 (96%) |  | 15 (100%) | 73 (95%) |  | 34 (89%) | 27 (100%) |  | 37 (90%) | 28 (100%) |  |
| Missing | 28 | 18 | 10 |  | 4 | 6 |  | 3 | 3 |  | 0 | 2 |  |
| **Potassium** | | | | | | | | | | | | | |
| H | 231 (49%) | 188 (49%) | 42 (49%) | 0.97 | 6 (46%) | 36 (50%) | 0.80 | 17 (47%) | 12 (52%) | 0.71 | 8 (32%) | 9 (47%) | 0.30 |
| L | 0 (0%) | 0 (0%) | 0 (0%) |  | 0 (0%) | 0 (0%) |  | 0 (0%) | 0 (0%) |  | 0 (0%) | 0 (0%) |  |
| N | 243 (51%) | 194 (51%) | 43 (51%) |  | 7 (54%) | 36 (50%) |  | 19 (53%) | 11 (48%) |  | 17 (68%) | 10 (53%) |  |
| Missing | 60 | 42 | 17 |  | 6 | 11 |  | 5 | 7 |  | 16 | 11 |  |
| **Chloride** | | | | | | | | | | | | | |
| H | 10 (2.0%) | 7 (1.7%) | 2 (2.2%) | **0.05** | 0 (0%) | 2 (2.6%) | 0.71 | 1 (2.6%) | 0 (0%) | 0.26 | 0 (0%) | 0 (0%) | 0.51 |
| L | 11 (2.2%) | 6 (1.5%) | 5 (5.4%) |  | 0 (0%) | 5 (6.5%) |  | 3 (7.9%) | 0 (0%) |  | 2 (4.9%) | 0 (0%) |  |
| N | 485 (96%) | 393 (97%) | 85 (92%) |  | 15 (100%) | 70 (91%) |  | 34 (89%) | 27 (100%) |  | 39 (95%) | 28 (100%) |  |
| Missing | 28 | 18 | 10 |  | 4 | 6 |  | 3 | 3 |  | 0 | 2 |  |
| **Bicarbonate** | | | | | | | | | | | | | |
| H | 25 (4.9%) | 18 (4.4%) | 7 (7.6%) | 0.30 | 2 (13%) | 5 (6.5%) | 0.24 | 4 (11%) | 2 (7.4%) | 0.82 | 2 (4.9%) | 1 (3.6%) | 1.00 |
| L | 49 (9.7%) | 37 (9.1%) | 10 (11%) |  | 0 (0%) | 10 (13%) |  | 4 (11%) | 4 (15%) |  | 1 (2.4%) | 1 (3.6%) |  |
| N | 432 (85%) | 351 (86%) | 75 (82%) |  | 13 (87%) | 62 (81%) |  | 30 (79%) | 21 (78%) |  | 38 (93%) | 26 (93%) |  |
| Missing | 28 | 18 | 10 |  | 4 | 6 |  | 3 | 3 |  | 0 | 2 |  |
| **Urea** | | | | | | | | | | | | | |
| H | 1 (0.2%) | 0 (0%) | 0 (0%) | 1.00 | 0 (0%) | 0 (0%) | 1.00 | 0 (0%) | 0 (0%) | 1.00 | 1 (2.4%) | 0 (0%) | 1.00 |
| L | 1 (0.2%) | 1 (0.2%) | 0 (0%) |  | 0 (0%) | 0 (0%) |  | 0 (0%) | 0 (0%) |  | 0 (0%) | 0 (0%) |  |
| N | 504(100%) | 405 (100%) | 92 (100%) |  | 15 (100%) | 77 (100%) |  | 38 (100%) | 27 (100%) |  | 40 (98%) | 28 (100%) |  |
| Missing | 28 | 18 | 10 |  | 4 | 6 |  | 3 | 3 |  | 0 | 2 |  |
| **Creatinine** | | | | | | | | | | | | | |
| H | 6 (1.2%) | 6 (1.5%) | 0 (0%) | 0.65 | 0 (0%) | 0 (0%) | 0.19 | 0 (0%) | 0 (0%) | 1.00 | 1 (2.4%) | 0 (0%) | 1.00 |
| L | 25 (4.9%) | 20 (4.9%) | 5 (5.4%) |  | 2 (13%) | 3 (3.9%) |  | 2 (5.3%) | 2 (7.4%) |  | 2 (4.9%) | 2 (7.1%) |  |
| N | 475 (94%) | 380 (94%) | 87 (95%) |  | 13 (87%) | 74 (96%) |  | 36 (95%) | 25 (93%) |  | 38 (93%) | 26 (93%) |  |
| Missing | 28 | 18 | 10 |  | 4 | 6 |  | 3 | 3 |  | 0 | 2 |  |
| **Bilirubin** | | | | | | | | | | | | | |
| H | 17 (3.4%) | 11 (2.7%) | 4 (4.3%) | 0.49 | 1 (6.7%) | 3 (3.9%) | 0.52 | 3 (7.9%) | 0 (0%) | 0.26 | 2 (4.9%) | 2 (7.1%) | 1.00 |
| L | 0 (0%) | 0 (0%) | 0 (0%) |  | 0 (0%) | 0 (0%) |  | 0 (0%) | 0 (0%) |  | 0 (0%) | 0 (0%) |  |
| N | 489 (97%) | 395 (97%) | 88 (96%) |  | 14 (93%) | 74 (96%) |  | 35 (92%) | 27 (100%) |  | 39 (95%) | 26 (93%) |  |
| Missing | 28 | 18 | 10 |  | 4 | 6 |  | 3 | 3 |  | 0 | 2 |  |
| **Alkaline phosphatase** | | | | | | | | | | | | | |
| H | 12 (2.4%) | 9 (2.2%) | 2 (2.2%) | 0.44 | 0 (0%) | 2 (2.6%) | 1.00 | 1 (2.6%) | 0 (0%) | 1.00 | 0 (0%) | 0 (0%) | 0.41 |
| L | 13 (2.6%) | 9 (2.2%) | 4 (4.3%) |  | 0 (0%) | 4 (5.2%) |  | 2 (5.3%) | 1 (3.7%) |  | 0 (0%) | 1 (3.6%) |  |
| N | 481 (95%) | 388 (96%) | 86 (93%) |  | 15 (100%) | 71 (92%) |  | 35 (92%) | 26 (96%) |  | 41 (100%) | 27 (96%) |  |
| Missing | 28 | 18 | 10 |  | 4 | 6 |  | 3 | 3 |  | 0 | 2 |  |
| **Aspartate transferase** | | | | | | | | | | | | | |
| H | 43 (8.8%) | 33 (8.4%) | 10 (12%) | 0.35 | 1 (7.1%) | 9 (12%) | 1.00 | 4 (11%) | 2 (8.3%) | 1.00 | 6 (15%) | 2 (8.0%) | 0.47 |
| L | 0 (0%) | 0 (0%) | 0 (0%) |  | 0 (0%) | 0 (0%) |  | 0 (0%) | 0 (0%) |  | 0 (0%) | 0 (0%) |  |
| N | 443 (91%) | 359 (92%) | 76 (88%) |  | 13 (93%) | 63 (88%) |  | 31 (89%) | 22 (92%) |  | 34 (85%) | 23 (92%) |  |
| Missing | 48 | 32 | 16 |  | 5 | 11 |  | 6 | 6 |  | 1 | 5 |  |
| **Alanine transferase** | | | | | | | | | | | | | |
| H | 73 (14%) | 62 (15%) | 11 (12%) | 0.81 | 1 (6.7%) | 10 (13%) | 0.74 | 5 (13%) | 4 (15%) | 0.69 | 7 (17%) | 5 (18%) | 1.00 |
| L | 7 (1.4%) | 6 (1.5%) | 1 (1.1%) |  | 0 (0%) | 1 (1.3%) |  | 0 (0%) | 1 (3.7%) |  | 1 (2.4%) | 0 (0%) |  |
| N | 426 (84%) | 338 (83%) | 80 (87%) |  | 14 (93%) | 66 (86%) |  | 33 (87%) | 22 (81%) |  | 33 (80%) | 23 (82%) |  |
| Missing | 28 | 18 | 10 |  | 4 | 6 |  | 3 | 3 |  | 0 | 2 |  |
| **LDH** | | | | | | | | | | | | | |
| H | 80 (16%) | 63 (16%) | 16 (18%) | 0.79 | 1 (6.7%) | 15 (20%) | 0.40 | 9 (25%) | 4 (15%) | 0.19 | 10 (24%) | 7 (26%) | 0.89 |
| L | 18 (3.6%) | 12 (3.0%) | 3 (3.4%) |  | 0 (0%) | 3 (4.1%) |  | 0 (0%) | 2 (7.7%) |  | 0 (0%) | 0 (0%) |  |
| N | 400 (80%) | 326 (81%) | 70 (79%) |  | 14 (93%) | 56 (76%) |  | 27 (75%) | 20 (77%) |  | 31 (76%) | 20 (74%) |  |
| Missing | 36 | 23 | 13 |  | 4 | 9 |  | 5 | 4 |  | 0 | 3 |  |
| **CK** | | | | | | | | | | | | | |
| H | 40 (7.9%) | 31 (7.6%) | 9 (9.8%) | 0.28 | 1 (6.7%) | 8 (10%) | 1.00 | 3 (7.9%) | 3 (11%) | 0.82 | 4 (9.8%) | 4 (15%) | 0.82 |
| L | 2 (0.4%) | 1 (0.2%) | 1 (1.1%) |  | 0  (0%) | 1 (1.3%) |  | 1 (2.6%) | 0 (0%) |  | 1 (2.4%) | 0 (0%) |  |
| N | 464 (92%) | 374 (92%) | 82 (89%) |  | 14 (93%) | 68 (88%) |  | 34 (89%) | 24 (89%) |  | 36 (88%) | 23 (85%) |  |
| Missing | 28 | 18 | 10 |  | 4 | 6 |  | 3 | 3 |  | 0 | 3 |  |
| **Gamma GT** | | | | | | | | | | | | | |
| H | 32 (6.3%) | 26 (6.4%) | 5 (5.4%) | 0.66 | 0 (0%) | 5 (6.5%) | 0.76 | 3 (7.9%) | 1 (3.7%) | 0.53 | 3 (7.3%) | 0 (0%) | 0.16 |
| L | 12 (2.4%) | 8 (2.0%) | 3 (3.3%) |  | 0 (0%) | 3 (3.9%) |  | 2 (5.3%) | 0 (0%) |  | 3 (7.3%) | 0 (0%) |  |
| N | 462 (91%) | 372 (92%) | 84 (91%) |  | 15 (100%) | 69 (90%) |  | 33 (87%) | 26 (96%) |  | 35 (85%) | 28 (100%) |  |
| Missing | 28 | 18 | 10 |  | 4 | 6 |  | 3 | 3 |  | 0 | 2 |  |
| **Total protein** | | | | | | | | | | | | | |
| H | 2 (0.4%) | 1 (0.2%) | 1 (1.1%) | 0.30 | 0 (0%) | 1 (1.3%) | 1.00 | 1 (2.6%) | 0 (0%) | 1.00 | 0 (0%) | 0 (0%) | 1.00 |
| L | 7 (1.4%) | 5 (1.2%) | 2 (2.2%) |  | 0 (0%) | 2 (2.6%) |  | 1 (2.6%) | 1 (3.7%) |  | 1 (2.4%) | 0 (0%) |  |
| N | 497 (98%) | 400 (99%) | 89 (97%) |  | 15 (100%) | 74 (96%) |  | 36 (95%) | 26 (96%) |  | 40 (98%) | 28 (100%) |  |
| Missing | 28 | 18 | 10 |  | 4 | 6 |  | 3 | 3 |  | 0 | 2 |  |
| **Albumin** | | | | | | | | | | | | | |
| H | 27 (5.3%) | 21 (5.2%) | 6 (6.5%) | 0.61 | 0 (0%) | 6 (7.8%) | 0.58 | 4 (11%) | 2 (7.4%) | 1.00 | 1 (2.4%) | 0 (0%) | 1.00 |
| L | 0 (0%) | 0 (0%) | 0 (0%) |  | 0 (0%) | 0 (0%) |  | 0 (0%) | 0 (0%) |  | 0 (0%) | 0 (0%) |  |
| N | 479 (95%) | 385 (95%) | 86 (93%) |  | 15 (100%) | 71 (92%) |  | 34 (89%) | 25 (93%) |  | 40 (98%) | 28 (100%) |  |
| Missing | 28 | 18 | 10 |  | 4 | 6 |  | 3 | 3 |  | 0 | 2 |  |
| **Globulin** | | | | | | | | | | | | | |
| H | 2 (0.4%) | 2 (0.5%) | 0 (0%) | 0.82 | 0 (0%) | 0 (0%) | 1.00 | 0 (0%) | 0 (0%) | 0.51 | 0 (0%) | 0 (0%) | 1.00 |
| L | 14 (2.8%) | 11 (2.7%) | 3 (3.3%) |  | 0 (0%) | 3 (3.9%) |  | 2 (5.3%) | 0 (0%) |  | 1 (2.4%) | 0 (0%) |  |
| N | 490 (97%) | 393 (97%) | 89 (97%) |  | 15 (100%) | 74 (96%) |  | 36 (95%) | 27 (100%) |  | 40 (98%) | 28 (100%) |  |
| Missing | 28 | 18 | 10 |  | 4 | 6 |  | 3 | 3 |  | 0 | 2 |  |
| **Calcium** | | | | | | | | | | | | | |
| H | 7 (1.4%) | 4 (1.0%) | 3 (3.3%) | 0.12 | 0 (0%) | 3 (3.9%) | 1.00 | 2 (5.3%) | 1 (3.7%) | 0.75 | 0 (0%) | 0 (0%) | 1.00 |
| L | 8 (1.6%) | 6 (1.5%) | 2 (2.2%) |  | 0 (0%) | 2 (2.6%) |  | 0 (0%) | 1 (3.7%) |  | 1 (2.4%) | 0 (0%) |  |
| N | 491 (97%) | 396 (98%) | 87 (95%) |  | 15 (100%) | 72 (94%) |  | 36 (95%) | 25 (93%) |  | 40 (98%) | 28 (100%) |  |
| Missing | 28 | 18 | 10 |  | 4 | 6 |  | 3 | 3 |  | 0 | 2 |  |
| **Magnesium** | | | | | | | | | | | | | |
| H | 2 (0.4%) | 2 (0.5%) | 0 (0%) | 0.21 | 0 (0%) | 0 (0%) | 1.00 | 0 (0%) | 0 (0%) | 1.00 | 0 (0%) | 0 (0%) | 1.00 |
| L | 1 (0.2%) | 0 (0%) | 1 (1.1%) |  | 0 (0%) | 1 (1.3%) |  | 1 (2.6%) | 0 (0%) |  | 1 (2.4%) | 0 (0%) |  |
| N | 503 (99%) | 404 (100%) | 91 (99%) |  | 15 (100%) | 76 (99%) |  | 37 (97%) | 27 (100%) |  | 40 (98%) | 28 (100%) |  |
| Missing | 28 | 18 | 10 |  | 4 | 6 |  | 3 | 3 |  | 0 | 2 |  |
| **Phosphate** | | | | | | | | | | | | | |
| H | 13 (2.6%) | 8 (2.0%) | 5 (5.4%) | 0.08 | 1 (6.7%) | 4 (5.2%) | 0.66 | 2 (5.3%) | 2 (7.4%) | 0.88 | 2 (4.9%) | 2 (7.1%) | 0.56 |
| L | 53 (10%) | 46 (11%) | 6 (6.5%) |  | 0 (0%) | 6 (7.8%) |  | 3 (7.9%) | 3 (11%) |  | 4 (9.8%) | 5 (18%) |  |
| N | 440 (87%) | 352 (87%) | 81 (88%) |  | 14 (93%) | 67 (87%) |  | 33 (87%) | 22 (81%) |  | 35 (85%) | 21 (75%) |  |
| Missing | 28 | 18 | 10 |  | 4 | 6 |  | 3 | 3 |  | 0 | 2 |  |
| **Uric acid** | | | | | | | | | | | | | |
| H | 29 (5.7%) | 22 (5.4%) | 7 (7.6%) | 0.71 | 3 (20%) | 4 (5.2%) | **0.03** | 1 (2.6%) | 4 (15%) | 0.21 | 1 (2.4%) | 4 (14%) | 0.11 |
| L | 59 (12%) | 48 (12%) | 10 (11%) |  | 3 (20%) | 7 (9.1%) |  | 4 (11%) | 2 (7.4%) |  | 7 (17%) | 2 (7.1%) |  |
| N | 418 (83%) | 336 (83%) | 75 (82%) |  | 9 (60%) | 66 (86%) |  | 33 (87%) | 21 (78%) |  | 33 (80%) | 22 (79%) |  |
| Missing | 28 | 18 | 10 |  | 4 | 6 |  | 3 | 3 |  | 0 | 2 |  |
| **Triglycerides** | | | | | | | | | | | | | |
| H | 26 (20%) | 20 (20%) | 4 (17%) | 1.00 | 0 (0%) | 4 (19%) | 1.00 | 3 (27%) | 1 (20%) | 1.00 | 3 (10%) | 5 (26%) | 0.24 |
| L | 0 (0%) | 0 (0%) | 0 (0%) |  | 0(0%) | 0 (0%) |  | 0 (0%) | 0 (0%) |  | 0 (0%) | 0 (0%) |  |
| N | 102 (80%) | 81 (80%) | 20 (83%) |  | 3 (100%) | 17 (81%) |  | 8 (73%) | 4 (80%) |  | 26 (90%) | 14 (74%) |  |
| Missing | 406 | 323 | 78 |  | 16 | 62 |  | 30 | 25 |  | 12 | 11 |  |
| **Fasting triglycerides** | | | | | | | | | | | | | |
| H | 44 (12%) | 39 (13%) | 5 (7.4%) | 0.21 | 1 (8.3%) | 4 (7.1%) | 1.00 | 2 (7.4%) | 1 (4.5%) | 1.00 | 1 (8.3%) | 0 (0%) | 1.00 |
| L | 0 (0%) | 0 (0%) | 0 (0%) |  | 0 (0%) | 0 (0%) |  | 0 (0%) | 0 (0%) |  | 0 (0%) | 0 (0%) |  |
| N | 334 (88%) | 266 (87%) | 63 (93%) |  | 11 (92%) | 52 (93%) |  | 25 (93%) | 21 (95%) |  | 11 (92%) | 9 (100%) |  |
| Missing | 156 | 119 | 34 |  | 7 | 27 |  | 14 | 8 |  | 29 | 21 |  |
| **Cholesterol** | | | | | | | | | | | | | |
| H | 68 (53%) | 54 (53%) | 13 (54%) | 0.95 | 1 (33%) | 12 (57%) | 0.58 | 5 (45%) | 3 (60%) | 1.00 | 10 (34%) | 8 (42%) | 0.86 |
| L | 0 (0%) | 0 (0%) | 0 (0%) |  | 0 (0%) | 0 (0%) |  | 0 (0%) | 0 (0%) |  | 1 (3.4%) | 0 (0%) |  |
| N | 60 (47%) | 47 (47%) | 11 (46%) |  | 2 (67%) | 9 (43%) |  | 6 (55%) | 2 (40%) |  | 18 (62%) | 11 (58%) |  |
| Missing | 406 | 323 | 78 |  | 16 | 62 |  | 30 | 25 |  | 12 | 11 |  |
| **Fasting cholesterol** | | | | | | | | | | | | | |
| H | 167 (44%) | 143 (47%) | 24 (35%) | 0.08 | 4 (33%) | 20 (36%) | 1.00 | 8 (30%) | 8 (36%) | 0.62 | 9 (75%) | 5 (56%) | 0.40 |
| L | 0 (0%) | 0 (0%) | 0 (0%) |  | 0 (0%) | 0 (0%) |  | 0 (0%) | 0 (0%) |  | 0 (0%) | 0 (0%) |  |
| N | 211 (56%) | 162 (53%) | 44 (65%) |  | 8 (67%) | 36 (64%) |  | 19 (70%) | 14 (64%) |  | 3 (25%) | 4 (44%) |  |
| Missing | 156 | 119 | 34 |  | 7 | 27 |  | 14 | 8 |  | 29 | 21 |  |
| **HDL cholesterol** | | | | | | | | | | | | | |
| H | 176 (35%) | 141 (35%) | 31 (34%) | 0.94 | 6 (40%) | 25 (32%) | 0.91 | 13 (34%) | 7 (26%) | 0.72 | 12 (29%) | 6 (21%) | 0.22 |
| L | 40 (7.9%) | 31 (7.6%) | 8 (8.7%) |  | 1 (6.7%) | 7 (9.1%) |  | 5 (13%) | 3 (11%) |  | 2 (4.9%) | 5 (18%) |  |
| N | 290 (57%) | 234 (58%) | 53 (58%) |  | 8 (53%) | 45 (58%) |  | 20 (53%) | 17 (63%) |  | 27 (66%) | 17 (61%) |  |
| Missing | 28 | 18 | 10 |  | 4 | 6 |  | 3 | 3 |  | 0 | 2 |  |
| **LDL cholesterol** | | | | | | | | | | | | | |
| H | 166 (33%) | 137 (34%) | 28 (31%) | 0.63 | 5 (36%) | 23 (31%) | 0.71 | 8 (22%) | 9 (35%) | 0.28 | 15 (37%) | 9 (32%) | 0.70 |
| L | 0 (0%) | 0 (0%) | 0 (0%) |  | 0 (0%) | 0 (0%) |  | 0 (0%) | 0 (0%) |  | 0 (0%) | 0 (0%) |  |
| N | 332 (67%) | 264 (66%) | 61 (69%) |  | 9 (64%) | 52 (69%) |  | 28 (78%) | 17 (65%) |  | 26 (63%) | 19 (68%) |  |
| Missing | 36 | 23 | 13 |  | 5 | 8 |  | 5 | 4 |  | 0 | 2 |  |
| **Iron** | | | | | | | | | | | | | |
| H | 24 (4.7%) | 19 (4.7%) | 4 (4.3%) | 1.00 | 0 (0%) | 4 (5.2%) | 1.00 | 3 (7.9%) | 1 (3.7%) | 0.78 | 1 (2.4%) | 0 (0%) | 1.00 |
| L | 10 (2.0%) | 8 (2.0%) | 2 (2.2%) |  | 0 (0%) | 2 (2.6%) |  | 1 (2.6%) | 0 (0%) |  | 2 (4.9%) | 2 (7.1%) |  |
| N | 472 (93%) | 379 (93%) | 86 (93%) |  | 15 (100%) | 71 (92%) |  | 34 (89%) | 26 (96%) |  | 38 (93%) | 26 (93%) |  |
| Missing | 28 | 18 | 10 |  | 4 | 6 |  | 3 | 3 |  | 0 | 2 |  |
| **TIBC** | | | | | | | | | | | | | |
| H | 19 (3.8%) | 15 (3.7%) | 3 (3.3%) | 1.00 | 0 (0%) | 3 (4.0%) | 1.00 | 2 (5.4%) | 0 (0%) | 0.51 | 1 (2.4%) | 0 (0%) | 0.64 |
| L | 1 (0.2%) | 1 (0.2%) | 0 (0%) |  | 0 (0%) | 0 (0%) |  | 0 (0%) | 0 (0%) |  | 0 (0%) | 1 (3.7%) |  |
| N | 479 (96%) | 385 (96%) | 87 (97%) |  | 15 (100%) | 72 (96%) |  | 35 (95%) | 26 (100%) |  | 40 (98%) | 26 (96%) |  |
| Missing | 35 | 23 | 12 |  | 4 | 8 |  | 4 | 4 |  | 0 | 3 |  |
| **Transferrin saturation** | | | | | | | | | | | | | |
| H | 9 (1.8%) | 8 (2.0%) | 1 (1.1%) | 0.64 | 0 (0%) | 1 (1.3%) | 1.00 | 1 (2.7%) | 0 (0%) | 0.86 | 0 (0%) | 0 (0%) | 1.00 |
| L | 78 (16%) | 60 (15%) | 17 (19%) |  | 3 (20%) | 14 (19%) |  | 7 (19%) | 6 (23%) |  | 6 (15%) | 3 (11%) |  |
| N | 412 (83%) | 333 (83%) | 72 (80%) |  | 12 (80%) | 60 (80%) |  | 29 (78%) | 20 (77%) |  | 35 (85%) | 24 (89%) |  |
| Missing | 35 | 23 | 12 |  | 4 | 8 |  | 4 | 4 |  | 0 | 3 |  |
| **CRP highly sensitive** | | | | | | | | | | | | | |
| H | 37 (7.3%) | 33 (8.1%) | 4 (4.3%) | 0.21 | 1 (6.7%) | 3 (3.9%) | 0.52 | 0 (0%) | 2 (7.4%) | 0.17 | 2 (4.9%) | 2 (7.1%) | 1.00 |
| L | 0 (0%) | 0 (0%) | 0 (0%) |  | 0 (0%) | 0 (0%) |  | 0(0%) | 0 (0%) |  | 0 (0%) | 0 (0%) |  |
| N | 468 (93%) | 372 (92%) | 88 (96%) |  | 14 (93%) | 74 (96%) |  | 38 (100%) | 25 (93%) |  | 39 (95%) | 26 (93%) |  |
| Missing | 29 | 19 | 10 |  | 4 | 6 |  | 3 | 3 |  | 0 | 2 |  |
| **Troponin I highly sensitive** | | | | | | | | | | | | | |
| H | 4 (0.9%) | 4 (1.1%) | 0 (0%) | 1.00 | 0 (0%) | 0 (0%) | 1.00 | 0 (0%) | 0 (0%) | 1.00 | 0 (0%) | 0 (0%) | 1.00 |
| L | 0 (0%) | 0 (0%) | 0 (0%) |  | 0 (0%) | 0 (0%) |  | 0 (0%) | 0 (0%) |  | 0 (0%) | 0 (0%) |  |
| N | 458(99%) | 368 (99%) | 83 (100%) |  | 18 (100%) | 65 (100%) |  | 33 (100%) | 24 (100%) |  | 32 (100%) | 24 (100%) |  |
| Missing | 72 | 52 | 19 |  | 1 | 18 |  | 8 | 6 |  | 9 | 6 |  |
| **Amylase** | | | | | | | | | | | | | |
| H | 34 (7.3%) | 23 (6.2%) | 8 (9.6%) | **0.03** | 2  (11%) | 6 (9.2%) | 0.73 | 3 (9.1%) | 3 (12%) | 0.84 | 3 (9.4%) | 4 (17%) | 0.82 |
| L | 10 (2.2%) | 5 (1.3%) | 5 (6.0%) |  | 0 (0%) | 5 (7.7%) |  | 1 (3.0%) | 1 (4.2%) |  | 1 (3.1%) | 0 (0%) |  |
| N | 418 (91%) | 344 (92%) | 70 (84%) |  | 16 (89%) | 54 (83%) |  | 29 (88%) | 20 (83%) |  | 28 (88%) | 20 (83%) |  |
| Missing | 72 | 52 | 19 |  | 1 | 18 |  | 8 | 6 |  | 9 | 6 |  |
| **Ferritin** | | | | | | | | | | | | | |
| H | 61 (13.2%) | 48 (13%) | 13 (16%) | 0.76 | 2 (11%) | 11 (17%) | 0.78 | 6 (18%) | 3 (12%) | 0.72 | 6 (19%) | 4 (17%) | 1.00 |
| L | 11 (2.4%) | 10 (2.7%) | 1 (1.2%) |  | 0 (0%) | 1 (1.5%) |  | 0 (0%) | 0 (0%) |  | 1 (3.1%) | 0 (0%) |  |
| N | 390 (84%) | 314 (84%) | 69 (83%) |  | 16 (89%) | 53 (82%) |  | 27 (82%) | 21 (88%) |  | 25 (78%) | 20 (83%) |  |
| Missing | 72 | 52 | 19 |  | 1 | 18 |  | 8 | 6 |  | 9 | 6 |  |
| **Lipase** | | | | | | | | | | | | | |
| H | 36 (7.7%) | 29 (7.7%) | 5 (5.9%) | 0.88 | 1 (5.6%) | 4 (6.0%) | 0.26 | 2 (5.9%) | 2 (8.0%) | 1.00 | 3 (9.4%) | 1 (4.2%) | 0.63 |
| L | 5 (1.1%) | 4 (1.1%) | 1 (1.2%) |  | 1 (5.6%) | 0 (0%) |  | 0 (0%) | 0 (0%) |  | 0 (0%) | 0 (0%) |  |
| N | 425 (91%) | 341 (91%) | 79 (93%) |  | 16 (89%) | 63 (94%) |  | 32 (94%) | 23 (92%) |  | 29 (91%) | 23 (96%) |  |
| Missing | 68 | 50 | 17 |  | 1 | 16 |  | 7 | 5 |  | 9 | 6 |  |
| **Thyroid stimulating hormone** | | | | | | | | | | | | | |
| H | 3 (0.6%) | 3 (0.8%) | 0 (0%) | 1.00 | 0 (0%) | 0 (0%) | 1.00 | 0 (0%) | 0 (0%) | 1.00 | 1 (3.1%) | 1 (4.2%) | 1.00 |
| L | 0 (0%) | 0 (0%) | 0 (0%) |  | 0 (0%) | 0 (0%) |  | 0 (0%) | 0 (0%) |  | 0 (0%) | 0 (0%) |  |
| N | 464 (99%) | 372 (99%) | 85 (100%) |  | 18 (100%) | 67 (100%) |  | 34 (100%) | 25 (100%) |  | 31 (97%) | 23 (96%) |  |
| Missing | 67 | 49 | 17 |  | 1 | 16 |  | 7 | 5 |  | 9 | 6 |  |
| **Testosterone** | | | | | | | | | | | | | |
| H | 19 (4.1%) | 17 (4.6%) | 1 (1.2%) | 0.30 | 0 (0%) | 1 (1.5%) | **0.01** | 0 (0%) | 0 (0%) | 1.00 | 0 (0%) | 0 (0%) | 1.00 |
| L | 9 (1.9%) | 6 (1.6%) | 3 (3.6%) |  | 3 (17%) | 0 (0%) |  | 1 (3.0%) | 1 (4.2%) |  | 2 (6.2%) | 2 (8.3%) |  |
| N | 434 (94%) | 349 (94%) | 79 (95%) |  | 15 (83%) | 64 (98%) |  | 32 (97%) | 23 (96%) |  | 30 (94%) | 22 (92%) |  |
| Missing | 72 | 52 | 19 |  | 1 | 18 |  | 8 | 6 |  | 9 | 6 |  |
| **Insulin** | | | | | | | | | | | | | |
| H | 41 (8.9%) | 32 (8.6%) | 8 (9.9%) | 0.59 | 2 (12%) | 6 (9.4%) | 1.00 | 3 (9.4%) | 4 (17%) | 0.68 | 3 (9.4%) | 3 (12%) | 1.00 |
| L | 10 (2.2%) | 7 (1.9%) | 3 (3.7%) |  | 0 (0%) | 3 (4.7%) |  | 1 (3.1%) | 0 (0%) |  | 0 (0%) | 0 (0%) |  |
| N | 408 (89%) | 332 (89%) | 70 (86%) |  | 15 (88%) | 55 (86%) |  | 28 (88%) | 19 (83%) |  | 29 (91%) | 21 (88%) |  |
| Missing | 75 | 53 | 21 |  | 2 | 19 |  | 9 | 7 |  | 9 | 6 |  |
| **C peptide** | | | | | | | | | | | | | |
| H | 19 (4.1%) | 16 (4.3%) | 2 (2.4%) | 0.75 | 1 (5.9%) | 1 (1.5%) | 0.37 | 2 (6.1%) | 0 (0%) | 0.51 | 3 (9.4%) | 3 (12%) | 1.00 |
| L | 0 (0%) | 0 (0%) | 0 (0%) |  | 0 (0%) | 0 (0%) |  | 0 (0%) | 0 (0%) |  | 0 (0%) | 0 (0%) |  |
| N | 443 (96%) | 357 (96%) | 80 (98%) |  | 16 (94%) | 64 (98%) |  | 31 (94%) | 23 (100%) |  | 29 (91%) | 21 (88%) |  |
| Missing | 72 | 51 | 20 |  | 2 | 18 |  | 8 | 7 |  | 9 | 6 |  |
| **NT-proBNP** | | | | | | | | | | | | | |
| H | 2 (0.4%) | 1 (0.3%) | 1 (1.2%) | 0.45 | 1 (5.6%) | 0 (0%) | 0.22 | 0 (0%) | 1 (4.2%) | 0.42 | 0 (0%) | 0 (0%) | 1.00 |
| L | 0 (0%) | 0 (0%) | 0 (0%) |  | 0 (0%) | 0 (0%) |  | 0 (0%) | 0 (0%) |  | 0 (0%) | 0 (0%) |  |
| N | 460 (99%) | 371 (99%) | 82 (99%) |  | 17 (94%) | 65 (100%) |  | 33 (100%) | 23 (96%) |  | 32 (100%) | 24 (100%) |  |
| Missing | 72 | 52 | 19 |  | 1 | 18 |  | 8 | 6 |  | 9 | 6 |  |

Values presented as count and %. Cells with red shading indicating significant differences

Abbreviations: H, high; L, Low; N, Normal range; HCT, haematocrit test; MCV, Mean corpuscular volume; MCH, mean corpuscular haemoglobin; MCHC, mean corpuscular haemoglobin concentration; RDW, red cell distribution width; MPV, mean platelet volume; ESR, erythrocyte sedimentation rate; LDH, Lactate dehydrogenase; CK, Creatine Kinase; HDL, high-density lipoprotein; LDL, low-density lipoprotein; TIBC, total iron-binding capacity; CRP, C-reactive protein; NT-proBNP, N-terminal pro B-type natriuretic peptide.

**Table S3. Detailed CMR findings in new onset cardiac impairment at 12 months.** Red cells indicate abnormal values

|  |  | **A** | **B** | **C** | **D** | **E** | **F** | **G** | **H** | **I** | **J** |
| --- | --- | --- | --- | --- | --- | --- | --- | --- | --- | --- | --- |
| **Field Strength** |  | 1.5T | 1.5T | 1.5T | 1.5T | 1.5T | 3T | 1.5T | 1.5T | 3T | 3T |
| **sex** |  | F | F | F | M | M | F | F | M | M | F |
| **Age range** |  | 56-60 | 41-45 | 46-50 | 51-55 | 66-70 | 46-50 | 46-50 | 56-60 | 30-35 | 30-35 |
| **Global T1** | baseline | 1018 | 968 | 976 | 985 | 952 | 1219 | 1001 | 942 | 1141 | 1182 |
|  | follow up | 1025 | 1018 | 1024 | 998 | 988 | 1278 | 998 | 934 | 1170 | 1238 |
| **≥ 3 elevated T1 segments** | baseline | No | No | No | No | No | No | No | No | No | No |
|  | follow up | Yes | Yes | Yes | Yes | Yes | Yes | No | No | No | Yes |
| **Global T2** | follow up | 48 | NA | 47 | 47 | 47 | 45 | NA | 46 | NA | NA |
| **≥ 3 elevated T2 segments** | follow up | No | No | No | No | No | Yes | No | No | No | No |
| Left end diastolic volume (mL) | baseline | 65 | 81 | 80 | 89 | 81 | 90 | 75 | 117 | 80 | 73 |
|  | follow up | 64 | 91 | 83 | 78 | 86 | 91 | 68 | 103 | 79 | 54 |
| Left end systolic volume (mL) | baseline | 27 | 29 | 34 | 35 | 28 | 34 | 29 | 49 | 34 | 28 |
|  | follow up | 23 | 31 | 31 | 36 | 33 | 38 | 30 | 49 | 35 | 27 |
| Left ejection fraction (%) | baseline | 58 | 64 | 58 | 61 | 65 | 63 | 62 | 58 | 57 | 62 |
|  | follow up | 64 | 66 | 63 | 55 | 61 | 58 | 56 | 53 | 55 | 51 |
| Left stroke volume (mL) | baseline | 38 | 52 | 46 | 54 | 53 | 57 | 47 | 68 | 46 | 45 |
|  | follow up | 41 | 60 | 52 | 42 | 53 | 53 | 38 | 54 | 44 | 27 |
| Left ventricular max wall thickness (mm) | baseline | 10 | 9 | 7 | 9 | 9 | 9 | 8 | 10 | 10 | 9 |
|  | follow up | 11 | 8 | 8 | 12 | 9 | 7 | 9 | 12 | 8 | 11 |
| Left ventricular muscle mass (mm) | baseline | 79 | 79 | 61 | 99 | 84 | 56 | 63 | 134 | 74 | 83 |
|  | follow up | 89 | 75 | 65 | 118 | 82 | 58 | 67 | 149 | 70 | 92 |
| Left global circumferential strain (%) | baseline | -20 | -25 | -24 | -23 | -21 | -20 | -24 | -19 | -20 | -23 |
|  | follow up | -21 | -25 | -24 | -19 | -20 | -22 | NA | -17 | -19 | -19 |
| Left global longitudinal strain (%) | baseline | -13 | -18 | -14 | -15 | -15 | -15 | -16 | -9 | -14 | -16 |
|  | follow up | -13 | -17 | -18 | -16 | -12 | -17 | NA | -10 | -13 | -14 |
| Right end diastolic volume (mL) | baseline | 68 | 79 | 80 | 86 | 94 | 90 | 77 | 132 | 75 | 77 |
|  | follow up | 62 | 81 | 78 | 56 | 98 | 92 | 71 | 130 | 86 | 56 |
| Right end systolic volume (mL) | baseline | 31 | 28 | 34 | 32 | 39 | 40 | 31 | 60 | 33 | 28 |
|  | follow up | 31 | 28 | 31 | 23 | 41 | 41 | 37 | 71 | 44 | 28 |
| Right ejection fraction (%) | baseline | 54 | 64 | 58 | 63 | 58 | 56 | 59 | 55 | 56 | 63 |
|  | follow up | 50 | 65 | 60 | 58 | 58 | 55 | 48 | 45 | 49 | 49 |
| Right stroke volume (mL) | baseline | 36 | 51 | 46 | 54 | 55 | 50 | 46 | 72 | 42 | 48 |
|  | follow up | 31 | 52 | 47 | 33 | 57 | 51 | 34 | 59 | 42 | 27 |
